# Role of sex hormones in CD4^+^ T-cell mediated rheumatoid arthritis pathology

**DOI:** 10.64898/2025.12.17.25342530

**Authors:** Rohila Jha, Shubham Kumar Shaw, Gargee Bhattacharya, Harapriya Behera, Chandrasekhar Pattanaik, Soumya Sengupta, Prakash K Barik, Jyoti R Parida, Prasanta Padhan, Satish Devadas

## Abstract

**Background:** An adverse female sex-bias exists across many autoimmune disorders, yet its underlying mechanisms, particularly the role of sex hormones, remains poorly understood. Furthermore, the physiological influence of sex hormones in regulating T cell function remains undefined. We examined for the critical role of estrogen and progesterone, in regulating CD4^+^ T cell responses, specifically with respect to inflammation and their bone erosion potential in RA.

**Methods:** Inflammatory markers, circulating antibodies, sex hormone receptors, ERα and PR levels were investigated in both RA patients and controls. Further, RA CD4^+^ T cells were stimulated in varying concentrations of estradiol and progesterone and assessed for modulation in cytokines, transcription factors, RANKL, and FasL expression. Subsequent *ex-vivo* studies were performed to examine the role of sex hormones in modulating T cell responses.

**Results:** RA patients displayed systemic inflammation and high circulating antibodies, with significantly higher expression in synovial fluid. Higher expression of ERα and PR was evinced on RA CD4^+^ T cells. Upon hormone stimulation, two cohorts of patients namely responders and non-responders were observed with respect to modulation in cytokines, transcription factors, RANKL, and FasL expression. Our *ex-vivo* Th1 and Th17 cells demonstrated that sex hormones play a physiological role in modulating inflammation and have bone erosion potential.

**Conclusion:** Our findings demonstrate the pivotal significance of sex hormones in modulating TCR responses, thereby regulating inflammation and bone erosion in RA pathology. Further dissection of TCR signaling pathways with respect to sex hormone stimulation may provide promising targets for therapeutic implications.

**Graphical abstract:** 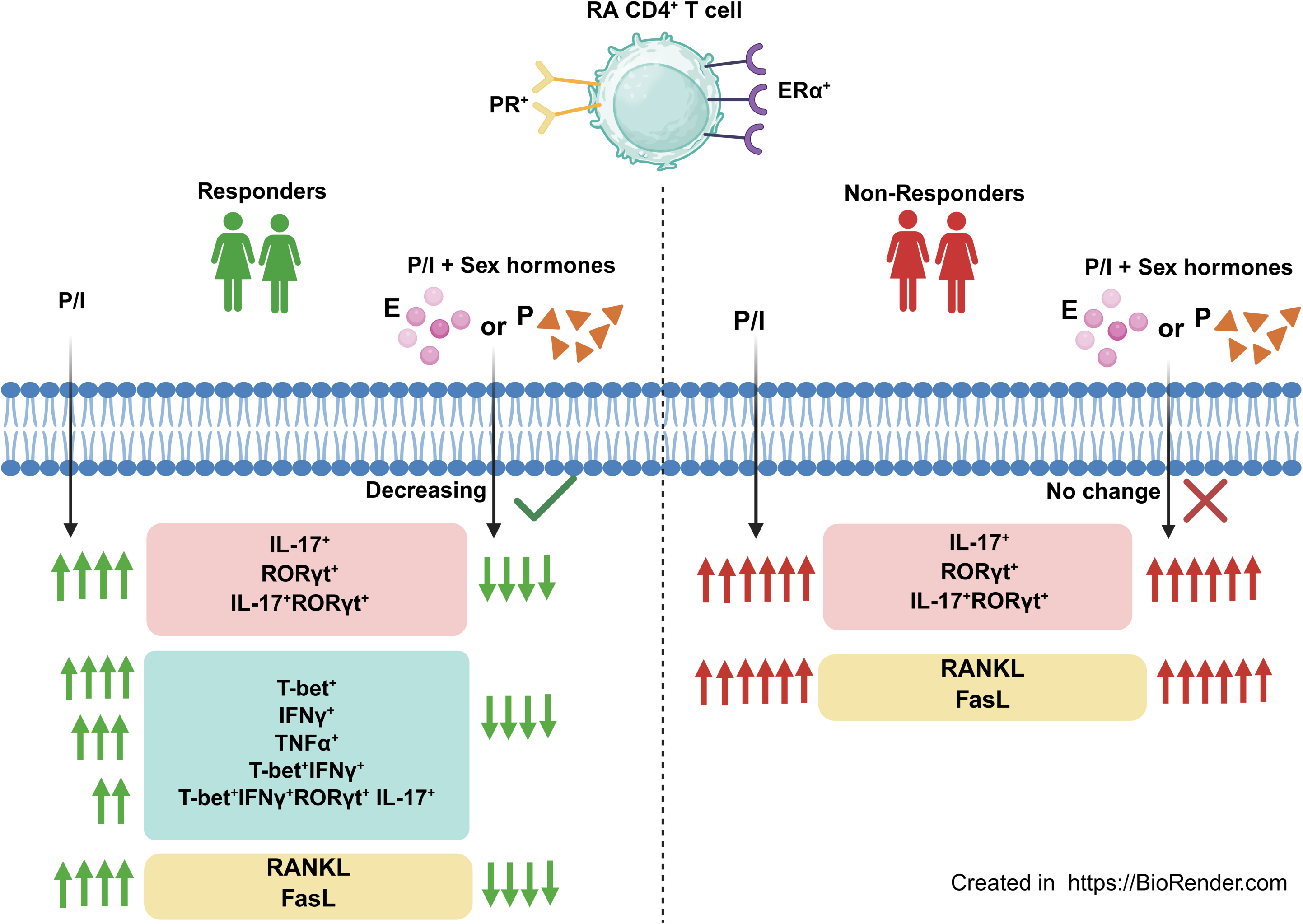

## 1. Introduction

T lymphocytes play a central role in adaptive immune system and they function via cognate antigen recognition and subsequent effector functions to eliminate pathogens and regulate immune tolerance [1]. However, T cells are also known to have a pivotal and well-defined role in driving the pathogenesis and exacerbation of autoimmune diseases [2]. Interestingly, a significant female preponderance is observed across most autoimmune disorders, with reported sex ratios ranging from approximately 2:1 to 20:1 [3–5]. This sexual dimorphism is observed not only in the incidence rates and prevalence but also in the severity of most autoimmune disorders, thereby implicating the possible role of sex hormones in contributing to disease pathogenesis. Nevertheless, the role of sex hormones specifically in the immune-endocrine interactions in autoimmune disorders such as Rheumatoid arthritis (RA) has not been fully elucidated and remains controversial. Interestingly, studies show the presence of sex hormone receptors such as ERa, ERβ, PR A/B, etc. on both innate and adaptive immune cells strongly suggesting their role in immune function [6, 7]. Sex hormone signaling has multifaceted roles such as differentiation and activation of immune cells, secretion of pro- and anti-inflammatory cytokines, thus maintaining immune homeostasis. These hormones are known to contribute sex-based differences in immune system, consequently predisposing women to autoimmune disorders. However, the dynamic physiological range of hormone levels across menstrual and reproductive cycles in women and significant disease heterogeneity collectively make it difficult to identify the mechanisms behind these hormones modulating immune functions.

Rheumatoid Arthritis (RA) is a chronic autoimmune rheumatic disorder, mainly characterized by sustained infiltration of immune cells into the synovial membrane leading to significant bone and cartilage erosion. Amongst multiple immune cells residing in synovial microenvironment, T helper cells play a central role in contributing to RA pathogenesis. These are aberrant CD4^+^ T cells which secretes various pro-inflammatory cytokines such as TNFα, IL-17, IL-1β and IL-6, stimulates B cells for autoantibodies production and promote RANKL-mediated bone erosion [8, 9]. Previous studies from our laboratory have reported this aberrant T cell to be a dual-positive “Th17Th1” phenotype with multi cytokine positivity *viz.* IFNγ^+^ IL-17^+^ TNFα^+^ GM-CSF^+^ RANKL^+^. Our studies also revealed the critical role of IL-21 and IL-23 in driving inflammation and bone erosion in RA [10, 11].

As evinced in other autoimmune disorders, the prevalence of RA is female sex-biased with a sex ratio of approximately 3:1. Furthermore, multiple observational studies suggest that female RA patients experience very severe and detrimental disease course compared to their male counterparts while limited studies have shown complex and contradictory roles of estrogen in RA [3, 12, 13]. Low levels of estrogen such as in post-menopausal state are associated with increased risk and exacerbated disease activity [14]. In contrast, high levels of estrogen in pregnancy are known to play a protective role [15]. However, progesterone plays an immunosuppressive role uniformly [16, 17]. Limited mice studies have examined the effects of estrogen or progesterone on the risk of RA [18–23]. The role of sex hormones in modulating immune responses in RA is a major focus of ongoing research. Furthermore, existing research characterizing the effects of sex hormones on CD4^+^ T cells in RA is limited and represents a critical gap in our understanding of disease’s pathogenesis.

Our work primarily focuses in establishing the role of sex hormones on CD4^+^ T cells with respect to inflammation and bone erosion. Our findings conclusively demonstrate higher expression of sex hormone receptors, ERα and PR on RA Th1, Th17 and dual positive subsets suggesting a link between endocrinology and immunology. Interestingly, we demonstrate two categories of RA patients *viz.* responders and non-responders, with respect to levels of inflammatory cytokines and associated transcription factor production upon TCR stimulation. Responders were able to downregulate cytokine and transcription factor expression while non-responders were unresponsive with all concentrations of both the hormones. Along with inflammation, we also demonstrate modulation in RANKL and FasL expression in RA patients. Taken together, our results indicate towards diverse processes being regulated by female sex hormones and understand its pivotal significance with respect to modulating TCR responses and in broad protection against autoimmunity.

## 2. Materials & Methods

### 2.1. Patients and controls

A total of 54 active RA patients were recruited in this study based on American College of Rheumatology/ European League Against Rheumatism 2010 (ACR/ EULAR) diagnostic criteria from the Out Patient Department of Odisha Arthritis and Rheumatology Centre (OARC), Bhubaneswar and Kalinga Institute of Medical Sciences (KIMS), Bhubaneswar between 2022 and 2025. The details of clinical parameters and medications taken by these patients are mentioned in **Table 1**. We also enrolled 25 gender- and age-matched healthy controls (HCs) defined in our inclusion criteria for controls. Informed consent was obtained from all participants. This study was done according to Helsinki declaration with the approval from the human ethics committee of the Institute of Life Sciences (HEC Ref No.: 76/HEC/18 and 132/ HEC/ 24).

**Table 1.**
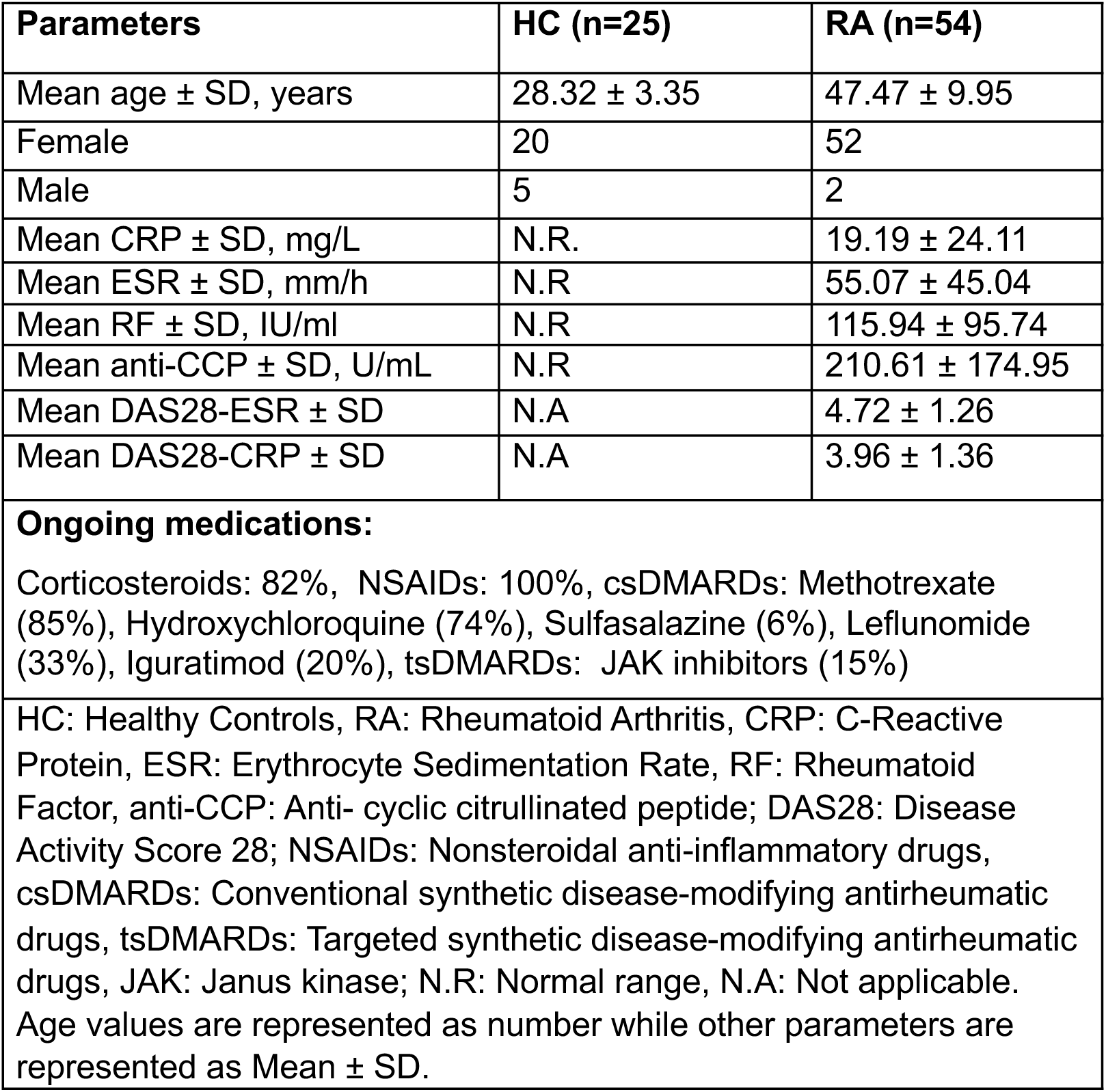
Clinical and demographic profiles of RA patients and healthy controls.

### 2.2. Plasma cytokines and circulating antibodies detection assay

Neat plasma derived from RA patients (n=15) and healthy controls (n=15) were run in duplicates to measure 46 different analytes including cytokines, chemokines, soluble receptors, and growth factors using a Human ProcartaPlex Mix & Match 46-Plex kit (Cat. No PPX-46-MX324DE, Invitrogen, Vienna, Austria), based on manufacturer’s instructions. Another set of neat plasma from same individuals, were analyzed by ProcartaPlex Human Antibody Isotyping Panels (Cat. No EPX070-10818-901, Invitrogen, Vienna, Austria). A set of synovial fluid and corresponding (paired) plasma of the same RA patients (n=9) and plasma derived from healthy controls (n=10) were run in duplicates to measure 48 different cytokines using a Bio-plex ProTM Human Cytokine Screening 48-Plex Panel (Cat. No #12007283). All the samples were acquired in Bio-Plex 200 system and the concentration of analytes were calculated using Bio-Plex manager software with a five-parameter (5PL) curve-fitting algorithm applied for standard curve calculation [24, 25].

### 2.3. Sex hormone receptor staining from PBMCs and SFMCs

5mL blood was collected from healthy volunteers and RA patients and PBMCs were isolated using Histopaque-1077 through density gradient centrifugation. Synovial fluid was collected from RA patients, SFMCs were isolated by treating synovial fluid with 300µg/mL Hyaluronidase for 20 minutes and then layered on Histopaque-1077 for density gradient centrifugation. For sex hormone receptors analyses on T helper subsets, the isolated cells were first stained with Zombie fixable violet dye kit (Bio Legend, San Diego, CA, USA) to exclude dead cells, followed by surface staining with fluorochrome tagged surface markers for 30 minutes and acquired in BD LSR Fortessa. The gating strategy is detailed in the supplementary figures **(Supplementary figure-1)**.

### 2.4. Activation of CD4^+^ T cells with sex hormones and staining for flow cytometry

5mL blood were collected from healthy volunteers and RA patients and PBMCs was isolated using Histopaque-1077 through density gradient centrifugation. CD4^+^ T cells were isolated from PBMCs by negative selection using Dyna beads (Invitrogen, MA, USA) according to manufacturer’s instructions. In brief, derived CD4^+^ T cells were seeded in flat bottomed plates at 1 million per mL cell density and were unstimulated without hormones or stimulated in presence or absence of either Estrogen (E2) or Progesterone (P4) across a range of three different concentrations in individual wells. The concentrations of E2 used were 1nM, 5nM and 10nM and the concentrations of P4 used was 16nM, 32nM and 64nM. The cells were further activated with PMA (25ng/mL), Ionomycin (500ng/mL) for 16 hours, with Brefeldin A (5µg/mL) added in the last 8 hours of stimulation. Stimulated cells were then stained with dead cell discrimination dye Zombie violet (Bio Legend, San Diego, CA, USA). For surface proteins staining, the cells were stained with marker specific antibodies. However, for transcription factor and cytokine staining, cells were first fixed with FOXP3 staining buffer set for 20 minutes. The fixed cells were then stained with specific fluorochrome-labelled antibodies for 30 minutes and acquired in BD LSR Fortessa. The gating strategy is detailed in the supplementary figures **(Supplementary figure-2,3)**.

### 2.5. *Ex-vivo* differentiation of human Th1 and Th17 cells

5mL blood was collected from healthy volunteers and PBMCs were isolated using Histopaque-1077 through density gradient centrifugation. CD4^+^ T cells were isolated from PBMCs by negative selection using Dyna beads (Invitrogen, MA, USA) according to manufacturer’s instructions. Cell purity was checked and ascertained to be higher than 85% (**Supplementary figure 4**). The isolated cells were cultured in RPMI 1640, supplemented with 10% fetal bovine serum of Australian origin, 100 U/mL Penicillin, 100 µg/mL Streptomycin, and 50 mM 2β-Mercapto ethanol (2β-ME). For Th1 differentiation, 1 million cells per mL density were plated on pre-coated αCD-3 (1 µg/mL) and αCD-28 (2 µg/mL) along with neutralizing antibodies αIL-4 (10 µg/mL), and cytokines IL-12 (10 ng/mL), IL-2 (100 IU/mL). The culture was activated for 5 days followed by resting for 2 days. For Th17 differentiation, 1 million cells per mL density were plated on pre-coated αCD-3 (1 µg/mL). αCD-28 (2 µg/mL) was added in soluble form along with neutralizing antibodies αIL-4 (10 µg/mL), αIFNγ (10 µg/mL) and cytokines, IL-1β (15 ng/mL), TGF-β (5 ng/mL), IL-21 (25 ng/mL), IL-23 (25 ng/mL) and IL-6 (25 ng/mL). After 7 days, both the cultures were washed with RPMI 1640, seeded in flat bottomed plates at 1 million per mL cell density and were unstimulated without hormones or stimulated in presence or absence of either Estrogen (E2) or Progesterone (P4) across a range of three different concentrations in individual wells. The concentrations of E2 used were 1nM, 5nM and 10nM and the concentrations of P4 used was 16nM, 32nM and 64nM. The cells were further activated with PMA (25ng/mL), Ionomycin (500ng/mL) for 16 hours, with Brefeldin A (5µg/mL) added in the last 8 hours of stimulation and used for subsequent experiments. The gating strategy is detailed in the supplementary figure **(Supplementary figure-5)**.

### 2.6. Statistics

Statistical analysis was performed using GraphPad Prism software, version 9.0.1. Data is presented as Mean ± standard error of mean (SEM). Mann-Whitney U test was used to compare statistics between healthy control and RA patients, including the cytokine and antibody profiling from bioplex. One way ANOVA test was used to compare cytokine expression in paired RA samples. Paired t-tests and one way ANOVA was used to compare cytokines, transcription factors, RANKL and FasL expression between E2 or P4 treated cells in RA patients and in terminally differentiated Th1 and Th17 cells.

## 3. Results

### 3.1. Dysregulated clinical parameters and disease scores in RA patients

The clinical characteristics of the patients showed higher C-Reactive Protein (CRP) and Erythrocyte Sedimentation Rate (ESR) levels, confirming active systemic inflammation (**Table 1**). All the patients were seropositive for both Rheumatoid factor (RF) and anti-cyclic citrullinated peptide (anti-CCP) antibodies, supporting diagnosis of RA. Corresponding disease activity scores were consistently high, with mean scores of DAS-28 ESR and DAS-28 CRP falling into the moderate to high disease activity category. Ongoing medications across the cohort showed that Nonsteroidal Anti-Inflammatory Drugs, were the most common class of medication prescribed, with all the patients receiving them, followed by Methotrexate, a Disease-Modifying Anti-Rheumatic Drug (85%) and corticosteroids (82%). This underscored a need to investigate their immune profile with respect to cytokines, circulating antibodies and T cell profile, all of which are discussed in detail.

### 3.2. RA patients display elevated cytokines and antibodies levels

To establish systemic inflammation in RA, we employed a multiplex assay system to quantify 46 analytes (comprising cytokines, chemokines, and soluble receptors) in RA patients’ plasma and observed significantly higher levels of 34 analytes in RA compared to HC. Based on their relative concentrations, analytes were categorized into high, medium, and low expression group and pro-inflammatory cytokines including IL-17, IL-6, IL-1β, IFN-γ, TNF-α, GM-CSF, IL-2, IL-22, TPO and VEGF-D were noted amongst high secretors apart from the chemokines MIP-1α in RA patients. Elevated levels of Th2 cytokines such as IL-10, IL-13, IL-9, IL-3, and IL-31 were also observed in RA plasma. Among the medium producers, we observed significant levels of IL-23, IL-21, IL-4, IL-5, LIF, IL-7, G-CSF, IL-27, IL-8, TSLP, IL-15 and IL-1α. In low producers, we observed IL-18, IFN-α, Eotaxin, CD62E, IL-37 and MCP-1 **(Fig- 1A)**. RA is characterized by synovial inflammation and therefore to correlate we analyzed cytokines in synovial fluid and plasma of paired RA samples. We observed differential expression and significantly higher levels of pro- and anti-inflammatory cytokines such as IL-6, IFN-γ, IP-10, LIF, MIG, SCF, IL-8, G-CSF, HGF, IL-2α, and IL-10 in RASF as opposed to RA plasma. Amongst the medium expressors were SCGF-β, RANTES, MIP-1α, IL-1Rα, IL-16, MCP-1, MCP-3, MCSF, SDF-1α, IL-15, IL-17A, Basic FGF and IL-12p40. In low expressors, we observed IFN-α2, IL-1β, IL-4 and TNF β **(Fig- 1B)**. To validate whether altered cytokine profile for sustained period could lead to skewed antibody profiles, we also investigated the levels of 7 different circulating antibodies and observed significantly high levels of IgG4, IgG1, IgA and IgG3 in RA patients as opposed to HCs **(Fig- 1C)**.

**Figure 1.**
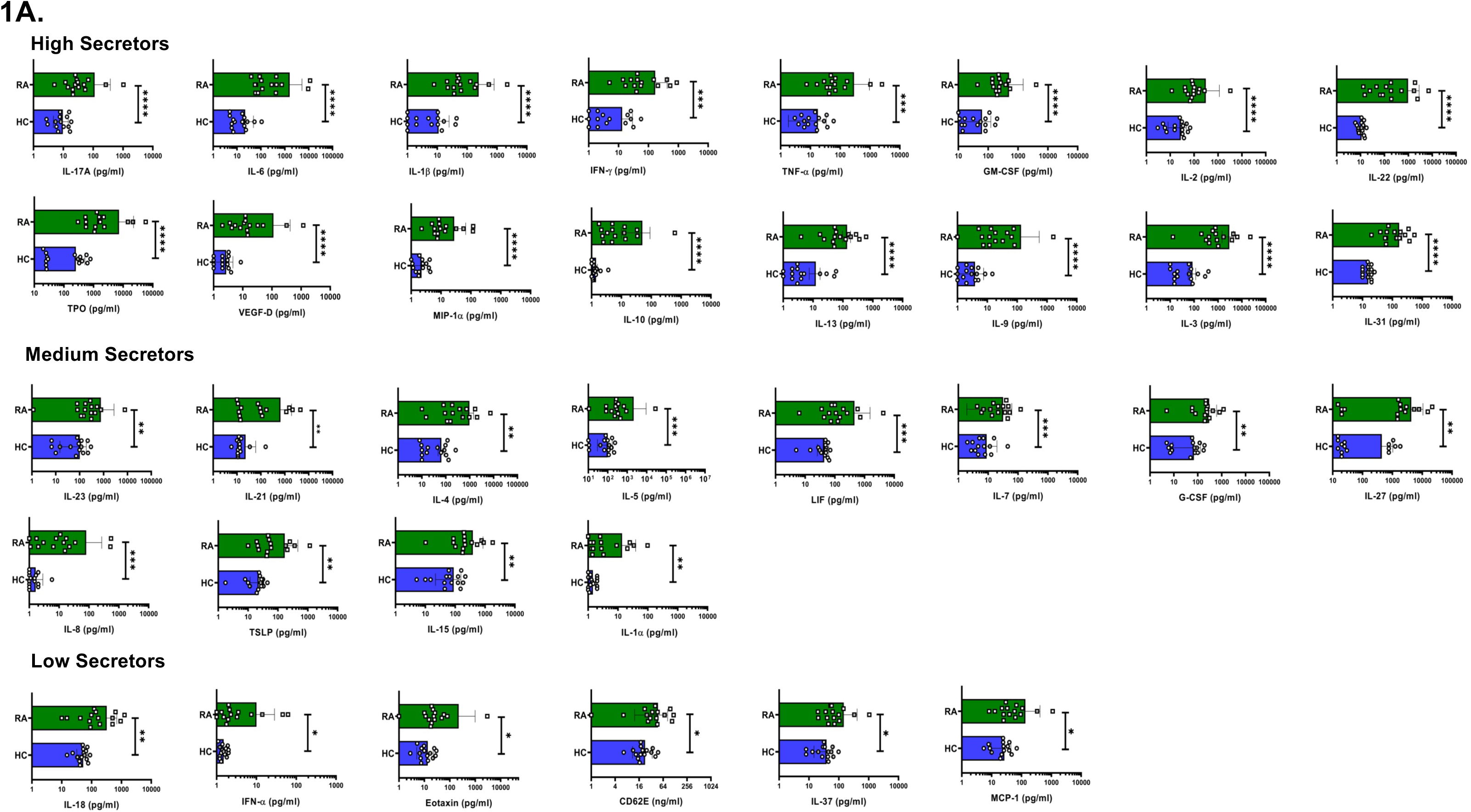
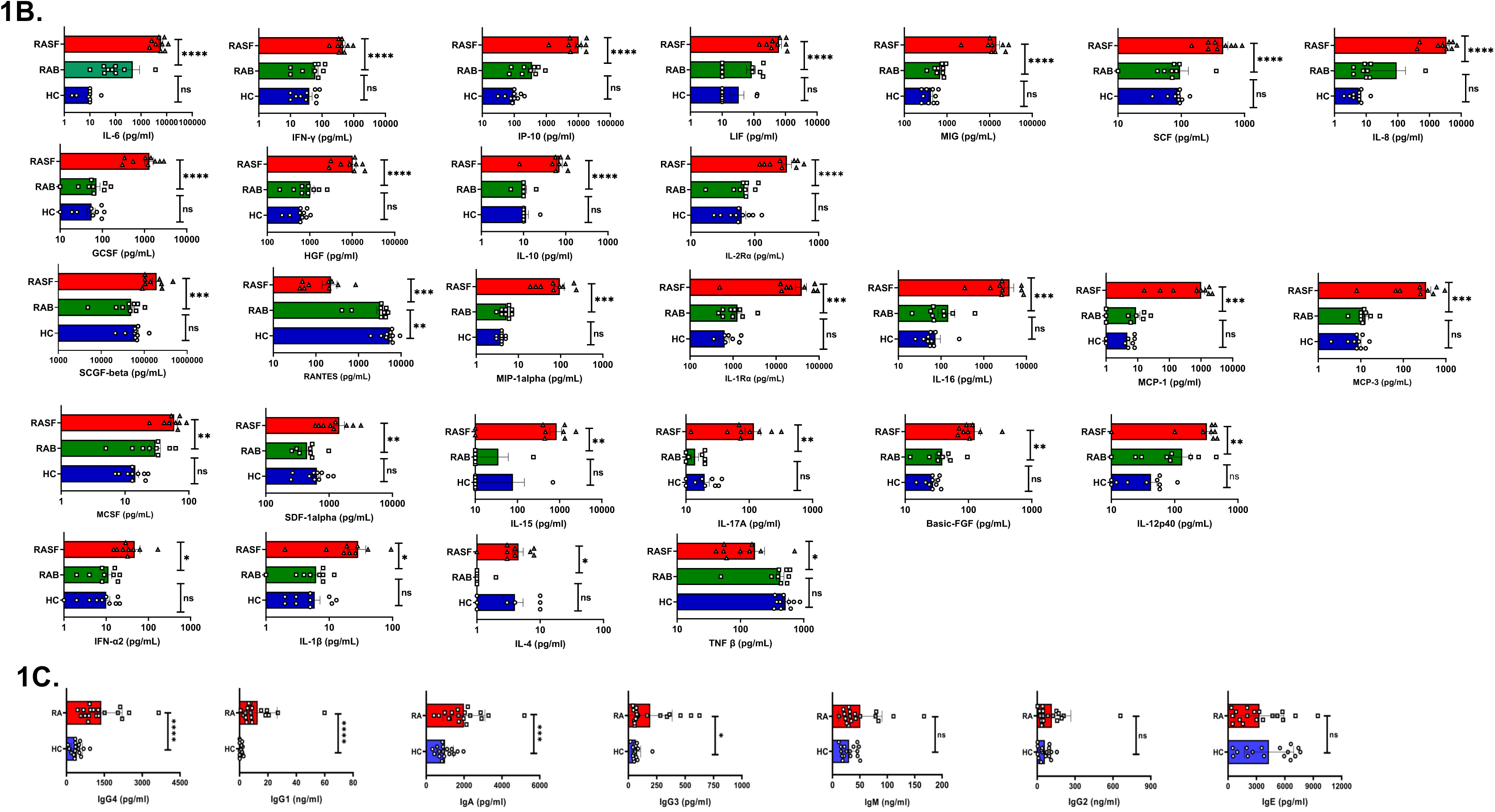
Plasma cytokine and antibody levels in RA patients and healthy controls. Representative figure showing differential skewed expression levels (high, medium & low) in RA patients’ plasma (n=15) vs healthy controls (n=15) are represented as graphical plots. **(A)**. Representative figure showing increased cytokine expression levels in RA patients’ synovial fluid (n=9) as compared to corresponding plasma vs healthy controls (n=9) are represented as graphical plots **(B)**. Amongst the circulating antibodies, IgG4, IgG1, IgA and IgG3 levels were elevated in RA patients (n=15) as compared to HC (n=15) **(C)**. Error bar indicates SEM. Mann–Whitney U Test was performed to compare between the two groups in figure 1A and 1B; one-way ANOVA test was performed to compare between the groups in figure 1C; p < 0.05 was considered statistically significant (*); p < 0.01 was considered to be very significant (**); p < 0.001 was considered to be highly significant (***); p < 0.0001 was considered extremely significant (****), and ns means non-significant.

### 3.3. RA CD4^+^ T cells display responder or non-responder cohorts with sex hormones

To assess the effects of exogenous sex hormones on TCR stimulated expression of various cytokines and transcription factors related to CD4^+^ T cells, a broad range of E2 or P4 concentrations based on normal ranges established in women were used. Interestingly, we observed two cohorts of patients, responders, and non-responders. In responder’s category, a significant decrease was observed in the stimulated response in cytokines such as IL-17 **(Fig- 2A & 2B)**, IFN-γ **(Fig- 2D & 2E)**, TNF-α **(Fig- 2F)** and transcription factors such as T-bet **(Fig- 2G)** and RORγt **(Fig- 2C)** expression with all the three concentrations of E2 or P4. Also, a three-category based response to TCR stimulation was observed at below 100, 100-300 and above 300% increase, with both the sex hormones in IFNγ **(Fig- 2D & 2E)** and TNFα **(Fig- 2F)** expression. In non-responder’s category, the basal inflammation was significantly high, observed for cytokines and transcription factors, such as IL-17 and RORγt **(Fig-2A to 2C)**. There was no reduction observed in their expression upon addition of any of the three concentrations of E2 or P4. Their stimulated response was much higher than responders. We also analyzed Th1, Th17 “Th17Th1” and “Th1Th17” dual positive subsets and observed a three-category based response viz. low, medium, and high responders in TCR stimulated response in Th1 (T-bet^+^ IFN-γ^+^) and Th17 Th1 (T-bet^+^ IFN-γ^+^ RORγt^+^ IL-17^+^) dual positive subsets with all the three concentrations of E2 and P4 **(Fig- 3A to 3C)**.

**Figure 2.**
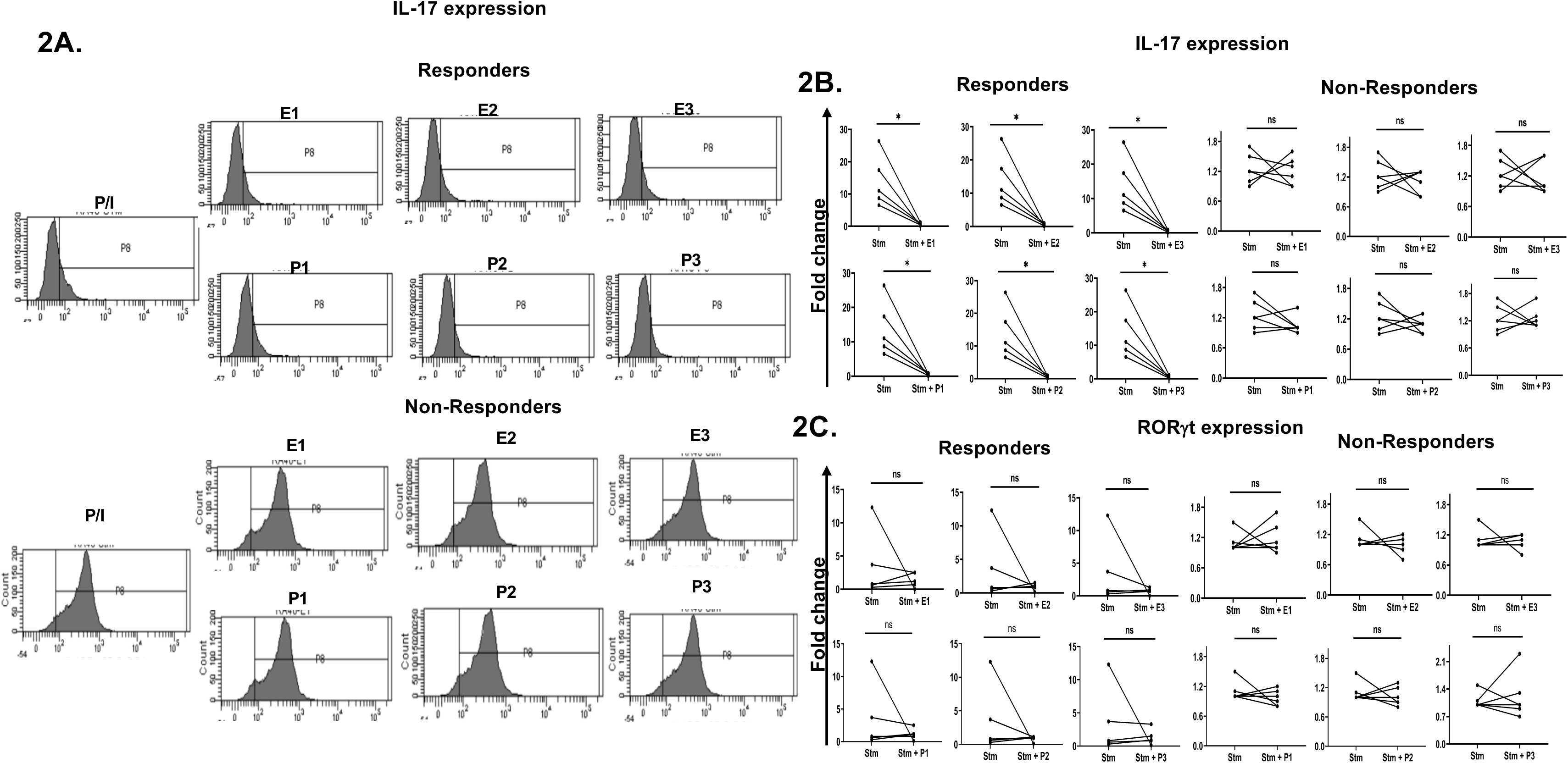
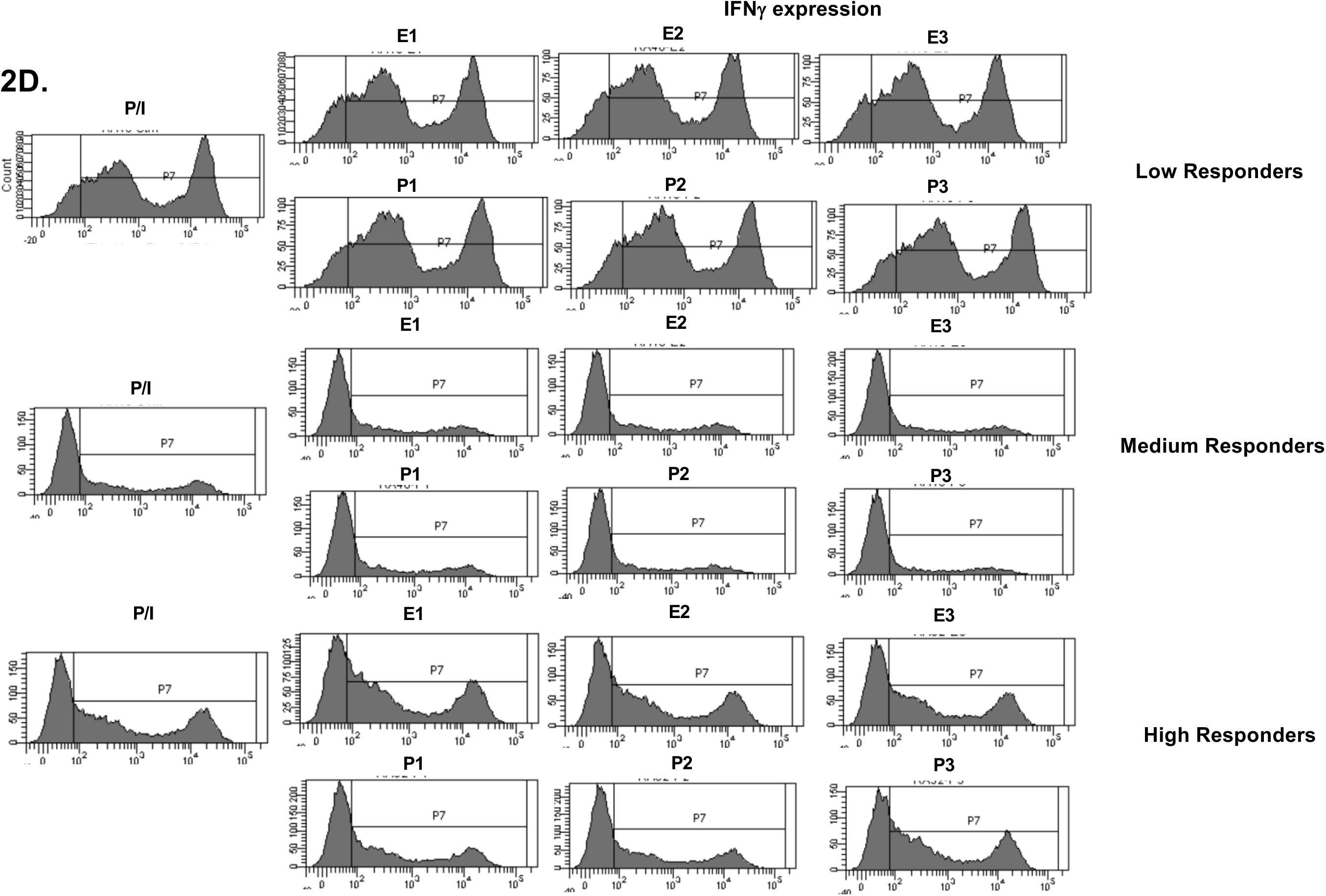
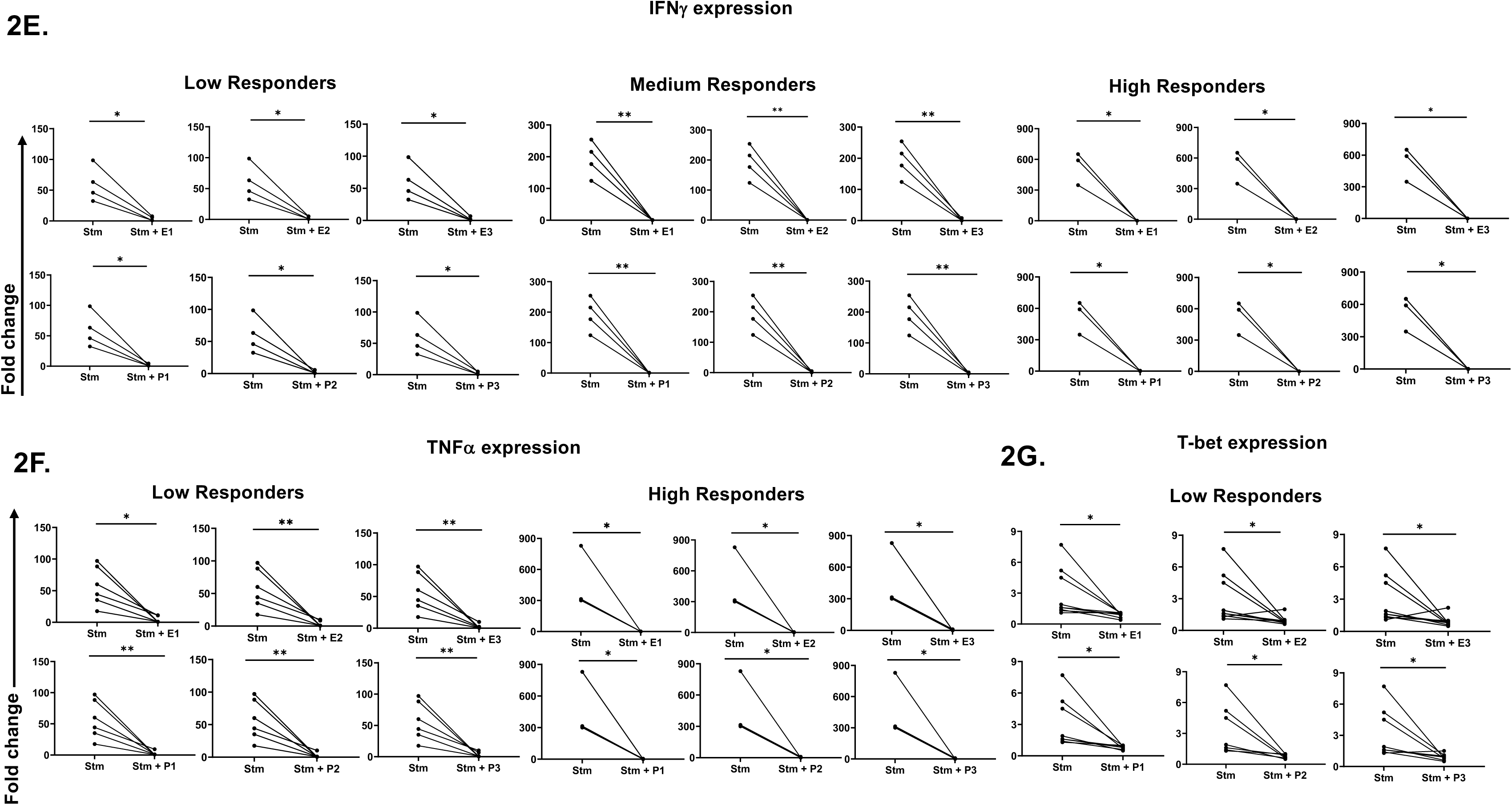
Effect of exogenous sex hormones (E2: 17β estradiol, P4: Progesterone) on expression of inflammatory cytokines and associated transcription factors in CD4^+^ T cells derived from RA PBMCs. Representative histogram plots **(A)** and cumulative graphical representation **(B, C)** showing responders and non-responders’ category of RA patients in presence of three different concentrations of E2 or P4 for IL-17 and RORγt expression. Representative histogram plots **(D)** and cumulative graphical representation showing low, medium, and high category-based response in the responders RA patients in presence of exogenous sex hormones of three different concentrations viz. E1, E2, E3, P1 P2 & P3 for IFNγ expression **(E)**, low and high responders for TNFα expression (**F)** and low responders for T-bet expression (**G)**. Paired t-test was performed to compare between the two groups, p < 0.05 was considered statistically significant (*); p < 0.01 was considered to be very significant (**); p < 0.001 was considered to be highly significant (***); p < 0.0001 was considered extremely significant (****), and ns means non-significant. Error bar represents SEM.

**Figure 3.**
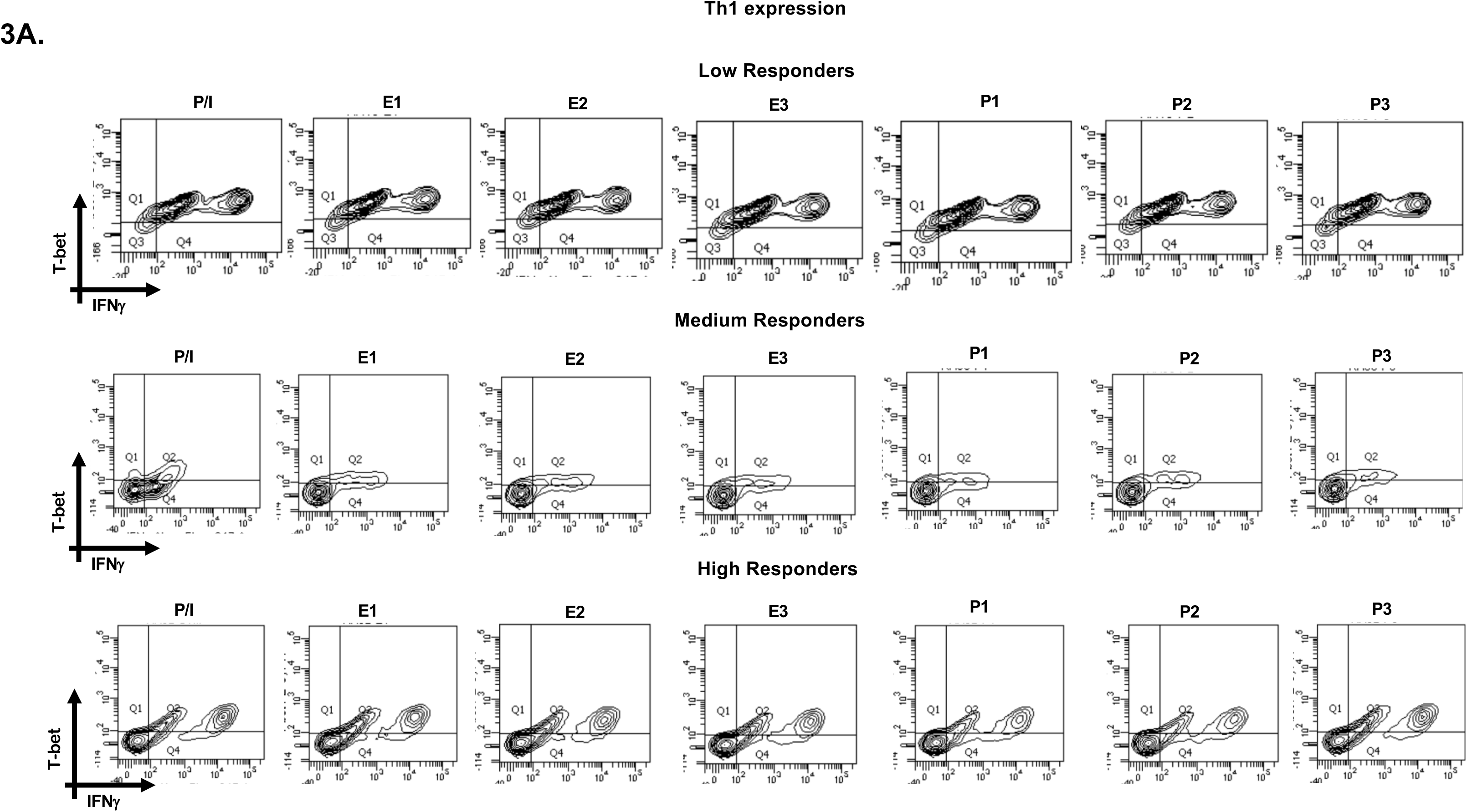
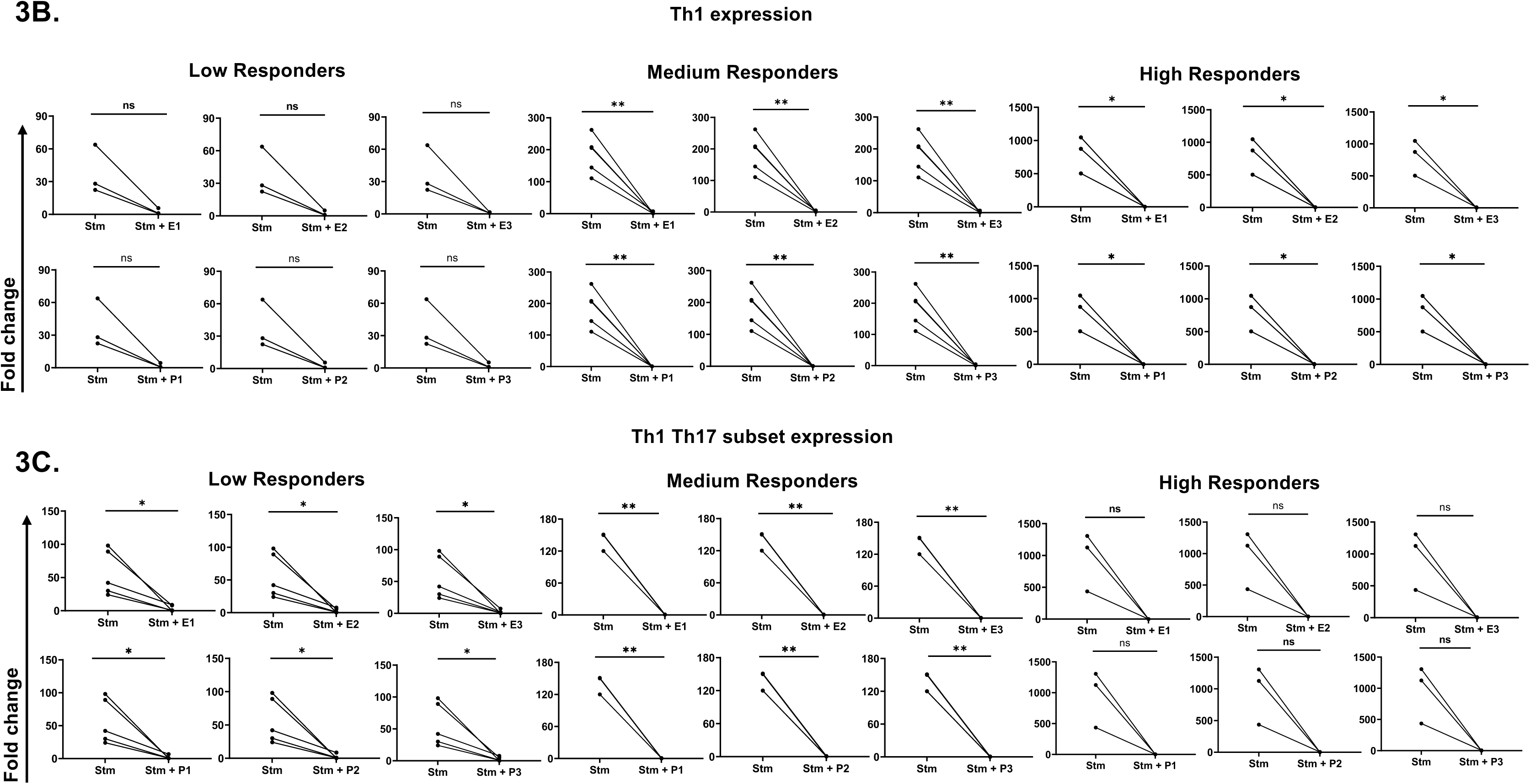
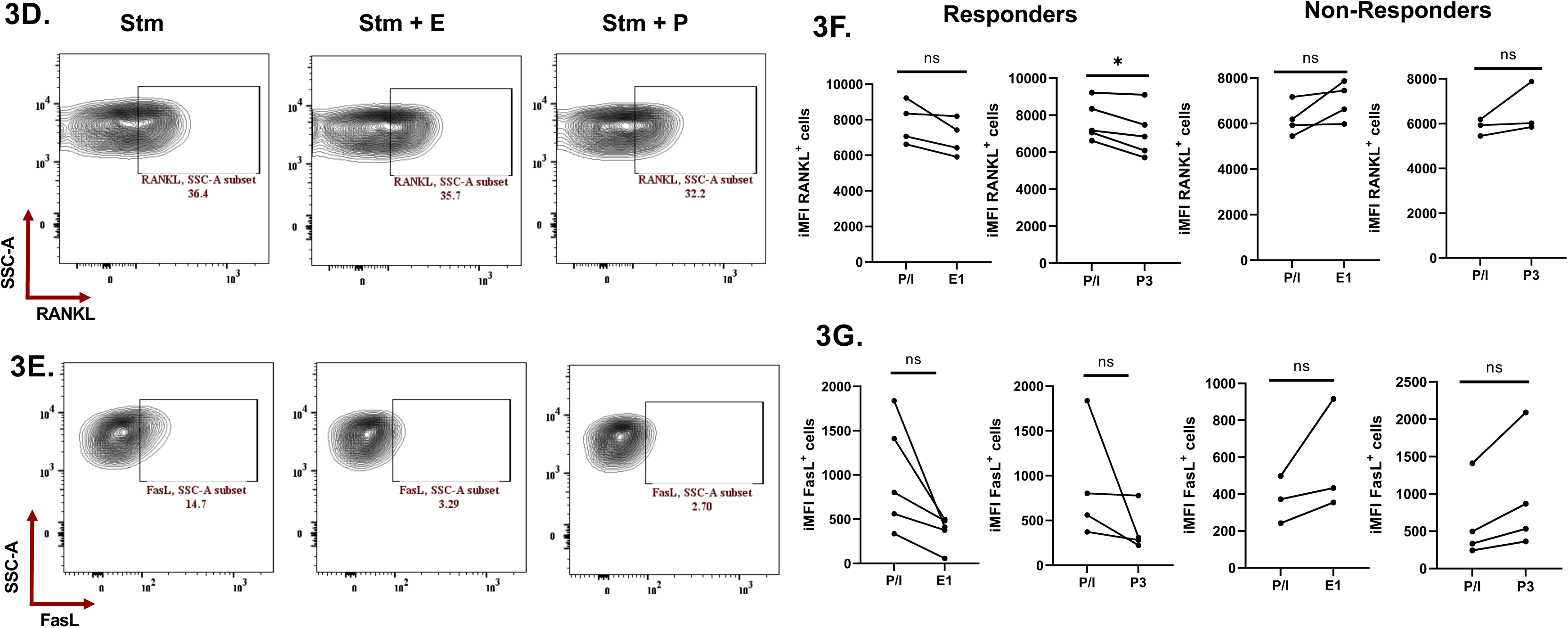
Effect of exogenous sex hormones (E2: 17β estradiol, P4: Progesterone) on Th1, Th17 and Th1 Th17 dual positive populations and RANKL & FasL proteins derived from RA PBMCs. Representative flow cytometry plots **(A)** and cumulative graphical representation **(B, C)** showing low, medium, and high category-based response in the responders RA patients in presence of exogenous sex hormones of three different concentrations viz. E1, E2, E3, P1 P2 & P3 for Th1 and Th1Th17 phenotype in RA patients. Representative flow cytometry plots **(D, E)** and cumulative graphical representation **(F, G)** showing responders and non-responders’ category for modulation with RANKL and FasL expression levels in presence of exogenous sex hormones. Paired t-test was performed to compare between the two groups, p < 0.05 was considered statistically significant (*); p < 0.01 was considered to be very significant (**); p < 0.001 was considered to be highly significant (***); p < 0.0001 was considered extremely significant (****), and ns means non-significant. Error bar represents SEM.

### 3.4. Sex hormones modulate RANKL and FasL expression in RA CD4^+^ T cells

In our next step, to assess the effects of exogenous sex hormones on proteins involved in osteoclastogenesis and tissue degradation in RA i.e. RANKL and FasL, we used E2 or P4 during TCR stimulation. We observed two cohorts of patients viz. responders and non-responders. There was a significant reduction in RANKL expression levels upon stimulation along with P4 treatment as compared to stimulated in the responder’s category **(Fig- 3D, 3F)**. Similarly, reduction was also observed in RANKL expression levels upon stimulation with E2 treatment as compared to stimulated alone in responder’s cohort **(Fig- 3D, 3F)**. In the non-responders’ cohort, no significant reduction was observed in the RANKL levels with E2 or P4 **(Fig- 3D, 3F)**. The expression levels of FasL also showed reduction with exogenous E2 or P4 in the responders’ category **(Fig- 3E, 3G)**. However, in the non-responders’ category there was no change in the expression of FasL levels when treated with E2 or P4 as compared to alone stimulated **(Fig- 3E, 3G)**.

### 3.5. Sex hormones modulate inflammatory cytokines, transcription factors, RANKL, and FasL in terminally differentiated human Th1 and Th17

To validate the role of sex hormones in modulating inflammatory cytokines and associated transcription factors expression and that the above results were not artifacts, we furthered our findings in *ex-vivo* differentiated human Th1 and Th17 cells. We restimulated the differentiated cells in the presence of different concentrations of E2 or P4 and analyzed the expression of cytokines and transcription factors. Our results demonstrated a significant downregulation of T-bet^+^, IFN-γ^+^ and T-bet^+^ IFN-γ^+^ population with all the three concentrations of E2 or P4 addition in Th1 cells **(Fig- 4A to 4D)**. Similarly, in Th17 cells we observed significant decrease in IL-17^+^ expression upon addition of all the three concentrations of E2 or P4. However, we did not observe any significant difference in RORγt^+^ and RORγt^+^IL-17^+^ population with E2 or P4 stimulation in Th17 cells **(Fig- 4E to 4H)**. Apart from inflammatory cytokines, only one concentration of exogenous estrogen i.e. E1 was able to modulate RANKL in Th1 cells, while other concentrations of sex hormones showed insignificant differences in RANKL expression **(Fig- 5A, 5C)**. Similarly, insignificant differences were observed in FasL expression with all the concentrations of E or P in Th1 cells **(Fig- 5B, 5D)**. In case of Th17 cells, RANKL expression displayed upregulation with P1 while all other hormone concentrations showed insignificant differences **(Fig- 5E, 5G).** Increase in expression, however statistically insignificant was observed in FasL expression in Th17 cells upon treatment with different concentrations of E or P **(Fig- 5F, 5H).**

**Figure 4.**
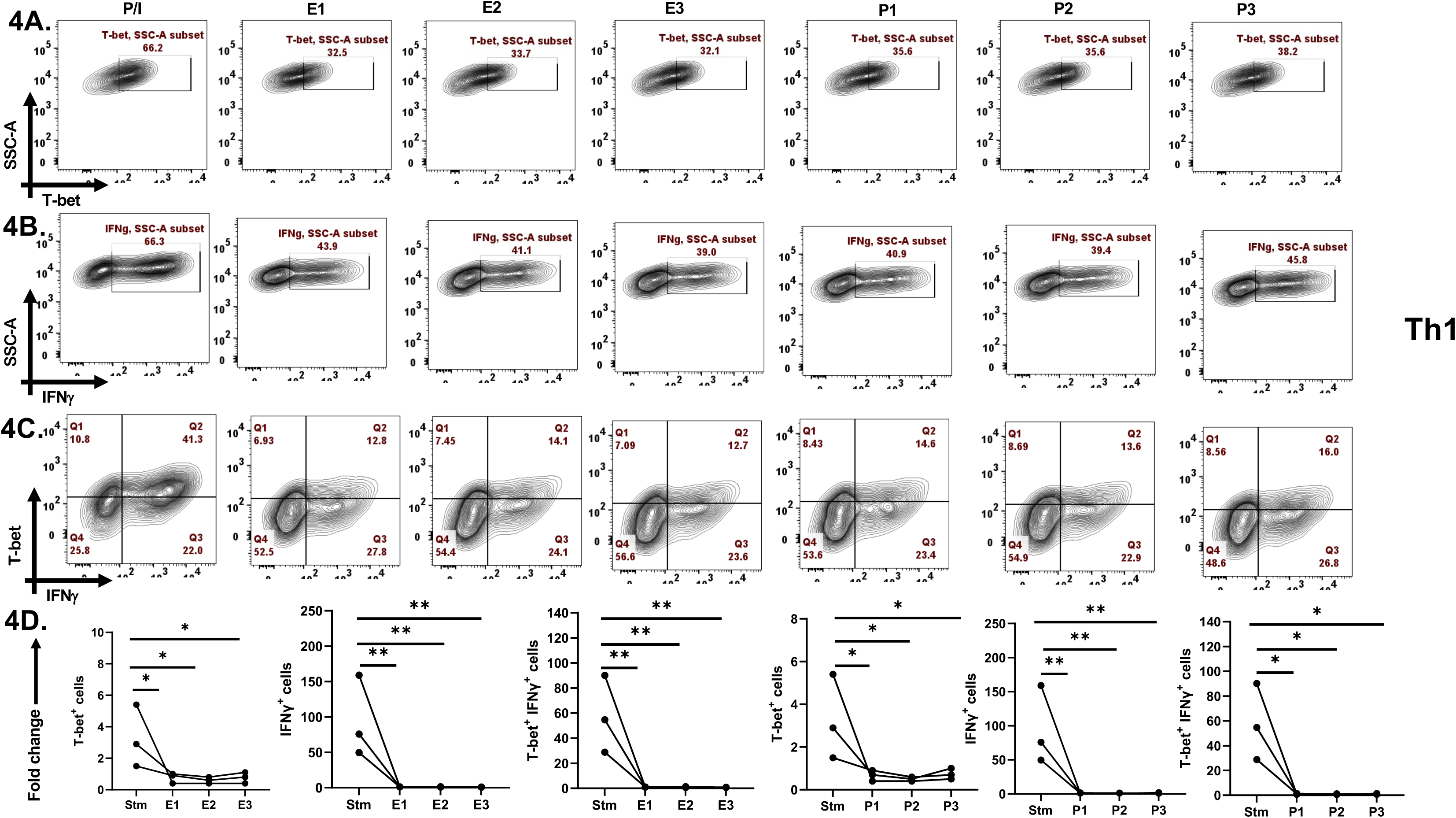
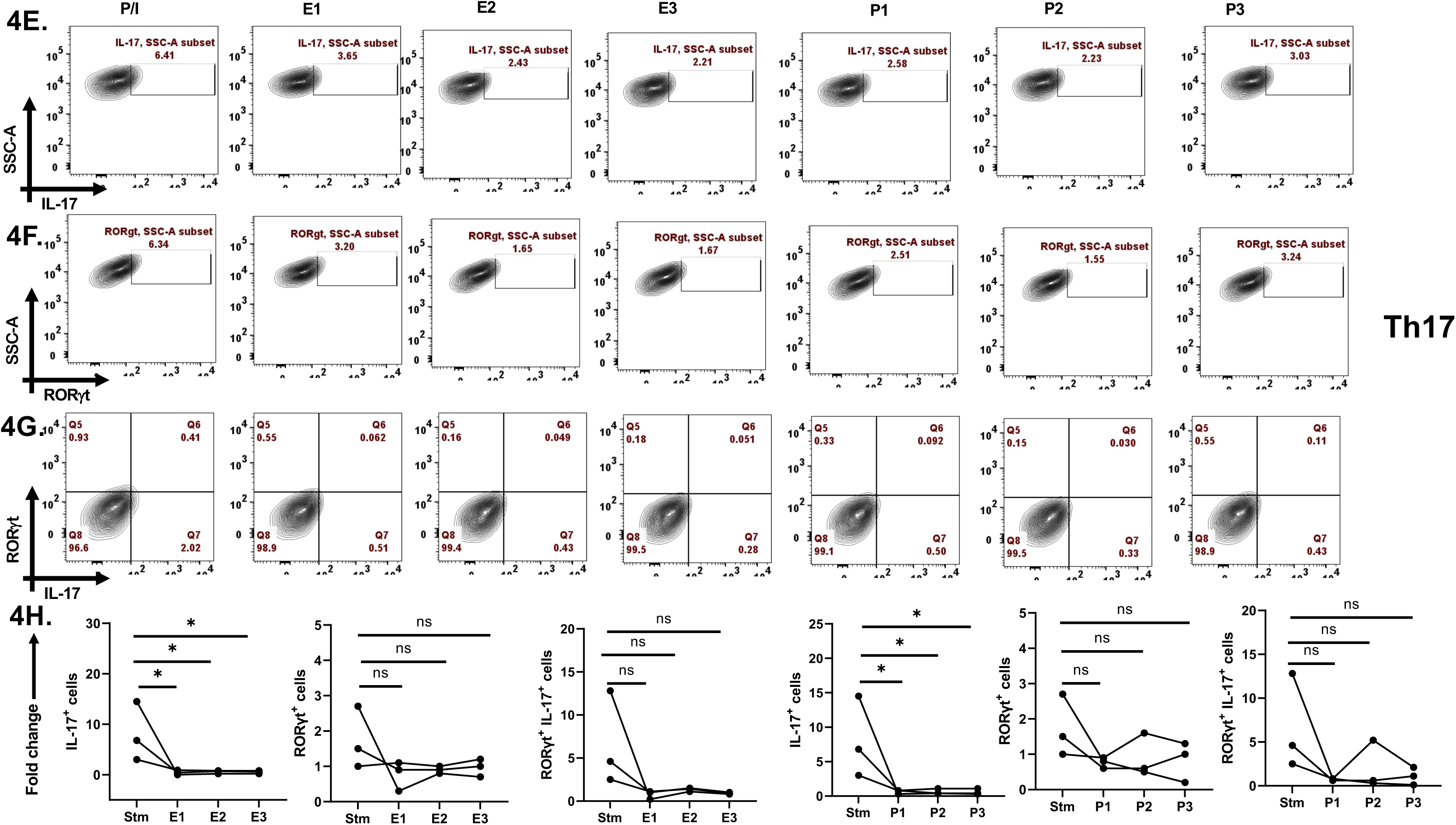
Effect of exogenous sex hormones (E2: 17β estradiol, P4: Progesterone) on cytokines and TFs expression in terminally differentiated human Th1 and Th17 cells. Negatively isolated CD4^+^ T cells from PBMCs were *ex-vivo* differentiated for 7 days into Th1 and Th17 phenotypes, following which they were reactivated with PMA/ Ion and different concentrations of E2 or P4 in individual wells for 16 hours. Representative flow cytometry plots **(A to C)** and cumulative graphical representation **(D)** showing modulation in T-bet+, IFNγ+ and T-bet^+^ IFNγ^+^ population in Th1 cells. Representative flow cytometry plots **(E to G)** and cumulative graphical representation **(H)** showing modulation in IL-17^+^, RORγt^+^ and RORγt^+^ IL-17^+^ population in Th17 cells. One-way ANOVA was performed to compare between the two groups, p < 0.05 was considered statistically significant (*); p < 0.01 was considered to be very significant (**); p < 0.001 was considered to be highly significant (***); p < 0.0001 was considered extremely significant (****), and ns means non-significant. Error bar represents SEM.

**Figure 5.**
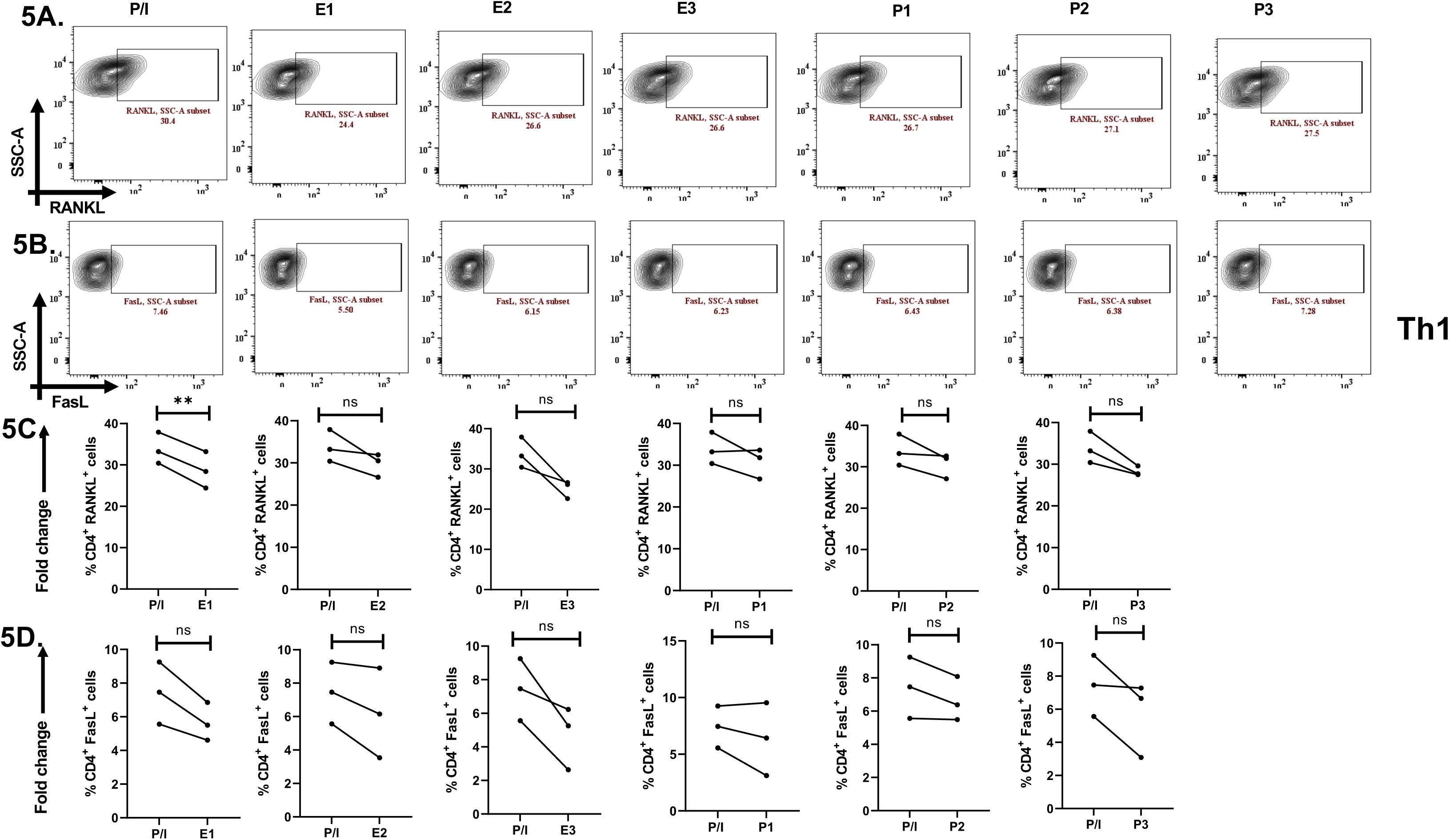
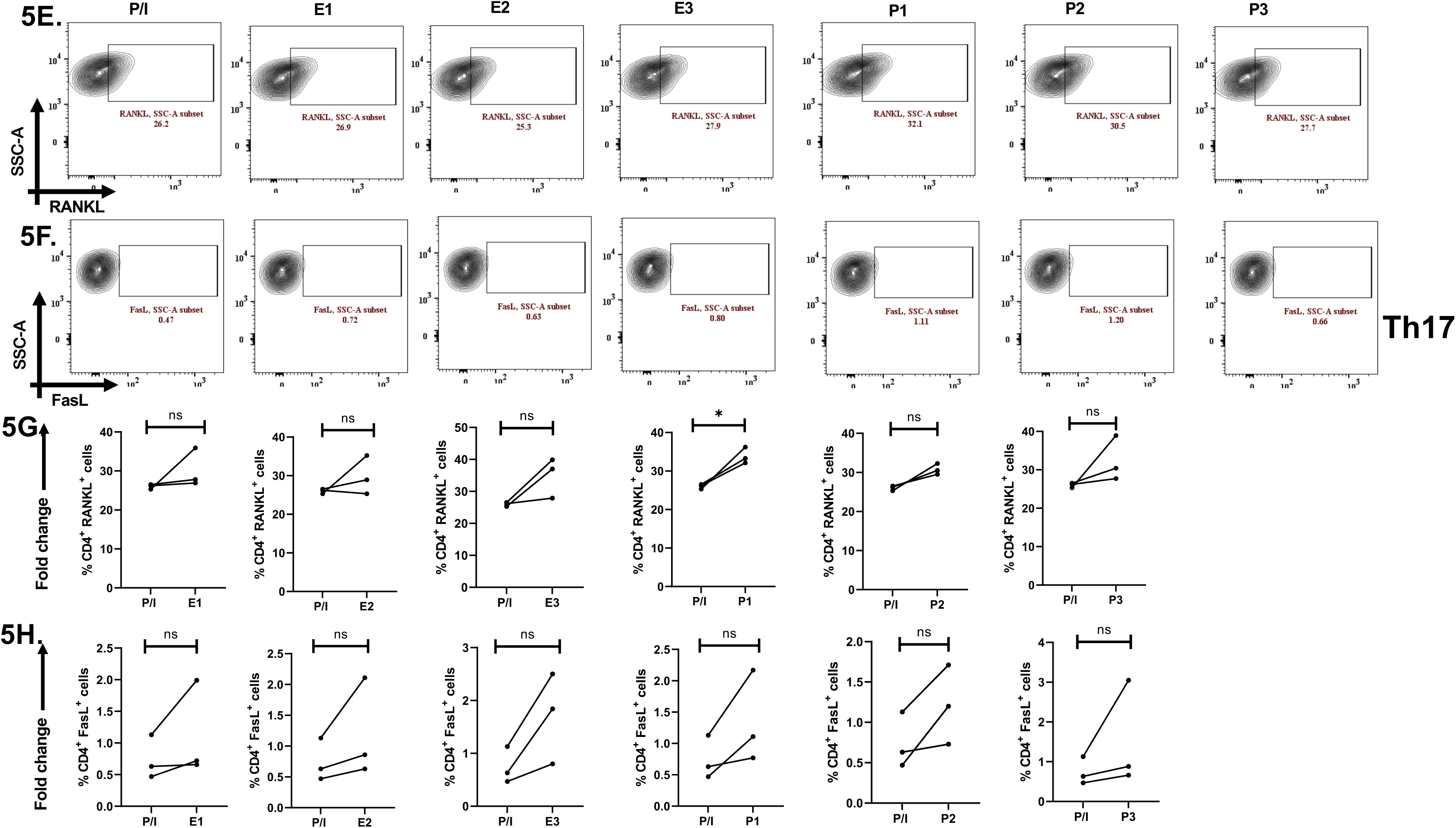
Effect of exogenous sex hormones (E2: 17β estradiol, P4: Progesterone) on RANKL and FasL proteins in terminally differentiated Th1 and Th17. Representative flow cytometry plots **(A, B)** and cumulative graphical representation **(C, D)** showing modulation in RANKL and FasL expression levels in presence of exogenous sex hormones in Th1 cells. Representative flow cytometry plots **(E, F)** and cumulative graphical representation **(G, H)** showing modulation in RANKL and FasL expression levels in presence of exogenous sex hormones in Th17 cells. Paired t-test was performed to compare between the two groups, p < 0.05 was considered statistically significant (*); p < 0.01 was considered to be very significant (**); p < 0.001 was considered to be highly significant (***); p < 0.0001 was considered extremely significant (****), and ns means non-significant. Error bar represents SEM.

## 4. Discussion

Although there is substantial evidence of adverse sex-biasness in autoimmune disorders such as RA, the definitive role of sex hormones in regulating T cell response remains inadequately understood [22]. Our work demonstrates an intriguing role for sex hormones in modulating CD4^+^ T cell immune responses, a major class of immune cells involved in RA pathogenesis, with regards to inflammation and bone erosion potential.

The clinical profile of recruited RA cohort (**Tab. 1**), characterized by elevated ESR and CRP levels, seropositive for both RF and anti-CCP antibodies, along with significantly high DAS28 scores demonstrates systemic inflammation and high disease activity. Such an active clinical state despite ongoing anti-rheumatic medications is a critical indicator of unresolved immune dysregulation in the patients. Furthermore, the notable female predominance in our patient cohort aligns with the strong sex-bias in RA prevalence, indicating sex hormones might play a role in regulating immune aberrancy. These clinical data provide a rationale for a deeper investigation to explore the role of immune dysregulation and inflammatory processes in RA.

To examine and establish inflammation in our RA patients we first analyzed for their cytokine levels in plasma to co relate with our clinical findings. Our multiplex data demonstrated elevated levels of pro-inflammatory cytokines in RA patients compared to healthy controls and for better analyses we classified them as high, medium, and low secretors. These analytes include inflammatory cytokines notably IL-17, IL-6, IL-1β, IFN-γ, TNF-α, GM-CSF, IL-15, IL-23, and IL-21 that are known to drive CD4^+^ T cell aberrancy, maintain and or exacerbate RA pathology. Higher expression of chemokines such as IL-8 and MCP-1 suggest constant recruitment and retention of other immune cells such as neutrophils, monocytes, and macrophages to maintain inflammation. Not surprisingly, the presence of significantly higher levels of Th2 cytokines such as IL-4, IL-5, IL-10, and IL-13 suggest a counter-response to inflammation. These results are consistent with prior studies suggesting concurrent presence of both inflammatory and anti-inflammatory cytokines in RA [10]. Further validation of our plasma findings was corroborated with significantly higher levels of a majority of cytokines and chemokines in synovial fluid of patients compared to their corresponding plasma. This clearly indicates that the synovial membrane is the major inflammatory site and immune cells within the joint actively produce and maintain high cytokine network and levels. The presence of cytokines within systemic circulation indicates an overflow or possible leak from the main source of joint(s) pathology. Apart from the cytokines, higher levels of circulating antibodies such as IgG4, IgG1, IgA and IgG3 in RA plasma suggest a direct co-relate with APC, B cells and T cells indicating sustained systemic inflammation for at least 4-6 weeks if not longer. The presence of significantly higher level of Th2 cytokines such as IL-4, IL-5, IL-10, and IL-13 was suggestive of B cell involvement and our isotype antibody clearly suggests this counterproductive immune response in RA patients [26, 27]. Taken together, the combined evidence from multiplexing and antibody isotyping data demonstrates a robust and sustained systemic inflammation in RA patients.

As our study focuses on the sex-biased CD4^+^ T cell responses in RA, we subsequently analysed for the expression of sex hormone receptors, ERα and PR and evinced a higher expression on RA Th1, Th17 and dual positive “Th17Th1” subsets **(Supplementary figure-6)**. This suggested a possible link between sex hormones and T cell aberrancy in RA pathology. Additionally, this suggests sex hormones upregulate their own receptor expression on T cells. To further identify their role in modulating T helper responses, we assessed their response when stimulated under varying comparable female physiological sex hormone concentration(s). Interestingly, our findings revealed two specific patient cohorts; *viz*. responders and non-responders. These cohorts showed differences in the threshold levels of inflammation, which was calculated as fold change in the iMFI levels of the stimulated alone and stimulated with hormones groups. Responders were able to show modulation with all the three concentrations of both estrogen or progesterone used as they demonstrated low or medium inflammatory cytokine profile. However, non-responders did not demonstrate modulation with any of the concentrations of estrogen or progesterone due to significantly much higher stimulated inflammation, strongly suggesting that the inflamed T cells were refractory. Interestingly, in the responders’ group both IL-17 and RORγt expression demonstrate significant reduction with E or P suggesting that sex hormones were able to modulate T cell responses. Cytokines such as IFNγ, TNFα & transcription factor such as T-bet showed three categories-based on response to TCR stimulation at <100, 100-300 and >300% fold change increase i.e. low, medium, and high responders. Also, the Th1 (T-bet^+^ IFNγ^+^) and Th17 Th1 dual positive (T-bet^+^ IFNγ^+^ RORγt^+^ IL-17^+^) phenotype showed response to all the three concentrations of both E and P in three categories viz. low, medium, and high responders indicating sex hormones modulate T cell profile based on the levels of basal inflammation. Crucially the non-responder group strongly suggested very significant inflammation and was refractory to sex hormones. Taken together, these findings strongly suggest the ability of sex hormones in modulating inflammatory status of T cells under a threshold and critically pointed also to a hyper inflamed non responding status.

Bone erosion and tissue degradation are another major characteristic of RA. Along with inflammation, our studies delineate the critical role of these hormones in modulating RANKL and FasL expression; key proteins mediating osteoclastogenesis and tissue degradation, respectively [28, 29]. Both these proteins are expressed on activated CD4^+^ T cells and are involved in RA pathogenesis [10, 30–32]. We demonstrate exogenous sex hormones modulating RANKL and FasL expression and observe the same two categories of patients as mentioned before *viz.* responders and non-responders, based on fold change in iMFI levels. Responders were able to downregulate RANKL with both estrogen and progesterone while non-responders, were unresponsive. Not surprisingly, reduction in FasL levels with E1 and P3 strongly indicated that sex hormones could modulate both bone and joint degrading proteins. These findings suggest that sex hormones could potentially modulate both FasL and RANK production and their TCR mediated upregulation and downstream signaling.

We furthered our understanding of the physiological role of sex hormones and their capability to modulate T cell responses by stimulating terminally differentiated Th1 and Th17 cells in presence of varying concentrations of sex hormones. Interestingly, we observed significant downregulation in Th1 and Th17 associated cytokines and transcription factors indicating estrogen and progesterone play a major role in immunomodulation. This again very strongly suggested a very intricate role *viz.* physiological one suggesting a fine control in bone morphogenesis and a modulatory one in inflammation when not too elevated.

Together these findings suggest a potential role of sex hormones especially estrogen and progesterone in shaping CD4^+^ T cell responses in RA patients via modulating cytokine levels, RANKL, and FasL levels. This underscores the need for further investigating the immune-endocrine interactions with respect to downstream mechanistic pathways involved in the same.

## Ethics Statement

The study was approved by the Institute Ethics Committee (IEC) / Institute Reference Board (IRB), Institute of Life Sciences with HEC reference number 76/HEC/18 & 132/ HEC/ 24 and was conducted according to the guidelines of Declaration of Helsinki.

## Data Availability Statement

All the data will be made available on request.

## Authors Contributions

Experimental design and conceptualization- SD and RJ

Sample collection, processing, and clinical scoring- JRP, PP, and PKB

Flow cytometry experiments- RJ, SKS, GB, and SS

Cytokine multiplexing and antibody isotyping experiments- RJ and SKS

Ex-vivo experiments- RJ, HB, and CP

Data collection and analysis- SD, RJ, and SKS

## Acknowledgements

We would like to thank Mr. Paritosh Nath, technical assistant in flow cytometry facility.

