## Supplementary file for "Role of sex hormones in CD4^+^ T-cell mediated rheumatoid arthritis pathology"

**Supplementary Figure 1.**

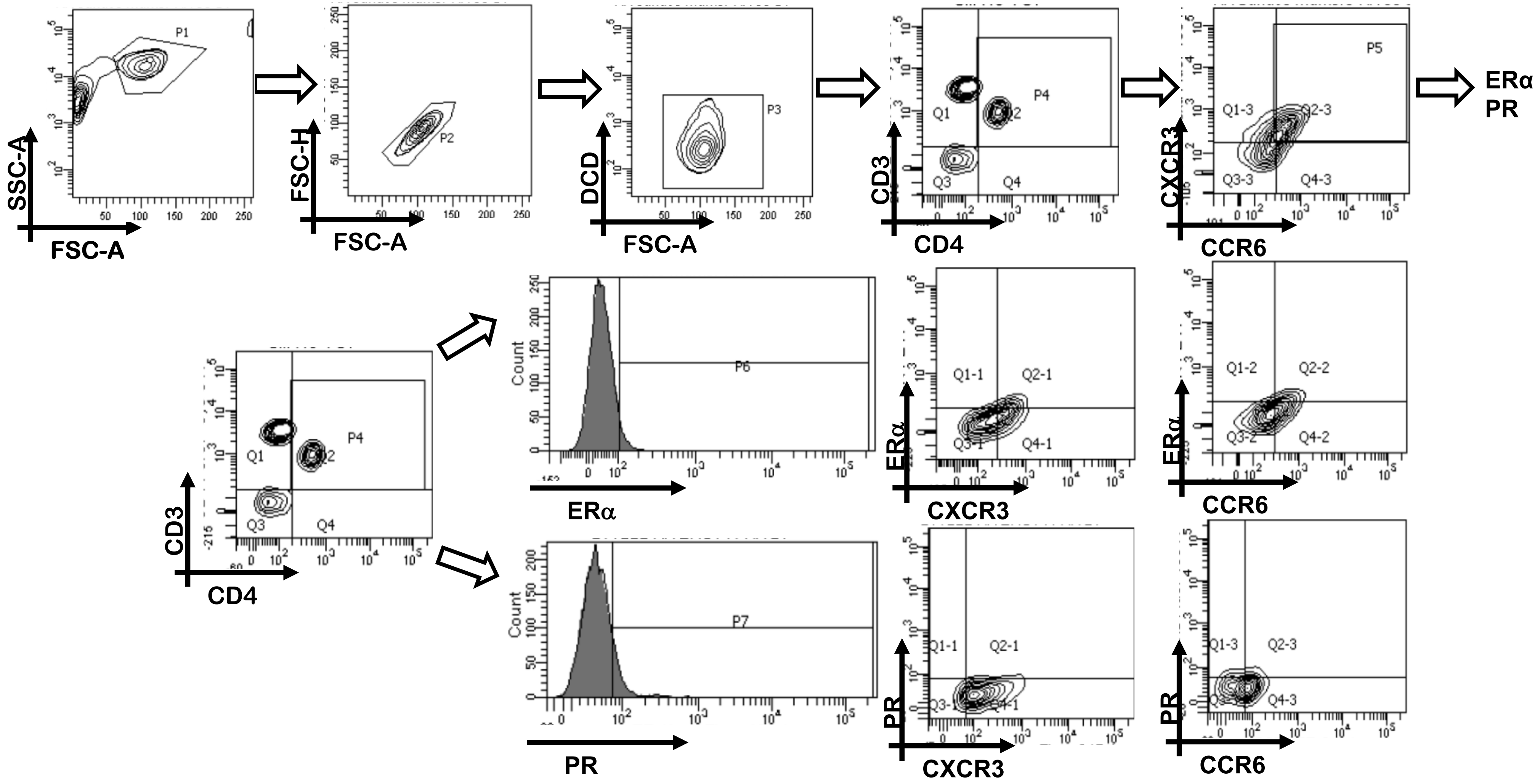


**Supplementary figure 1. Gating strategy for sex hormone receptors, ERα and PR expression in RA and HC CD4$^+$ T cells.** Representative flow cytometry plots shows lymphocyte population is gated first, followed by singlet population. Live cells are gated using Zombie violet dye. The dual positive CD3$^+$ CD4$^+$ T cells are selected to analyze for ERα and PR expression. Next, T-helper subset analysis was done based on surface marker CXCR3 for Th1 and CCR6 for Th17. ERα and PR was then analyzed on these T-helper subsets. Also, dual positive CXCR3$^+$CCR6$^+$ compartment was analyzed for ERα and PR expression.

**Supplementary Figure 2.**

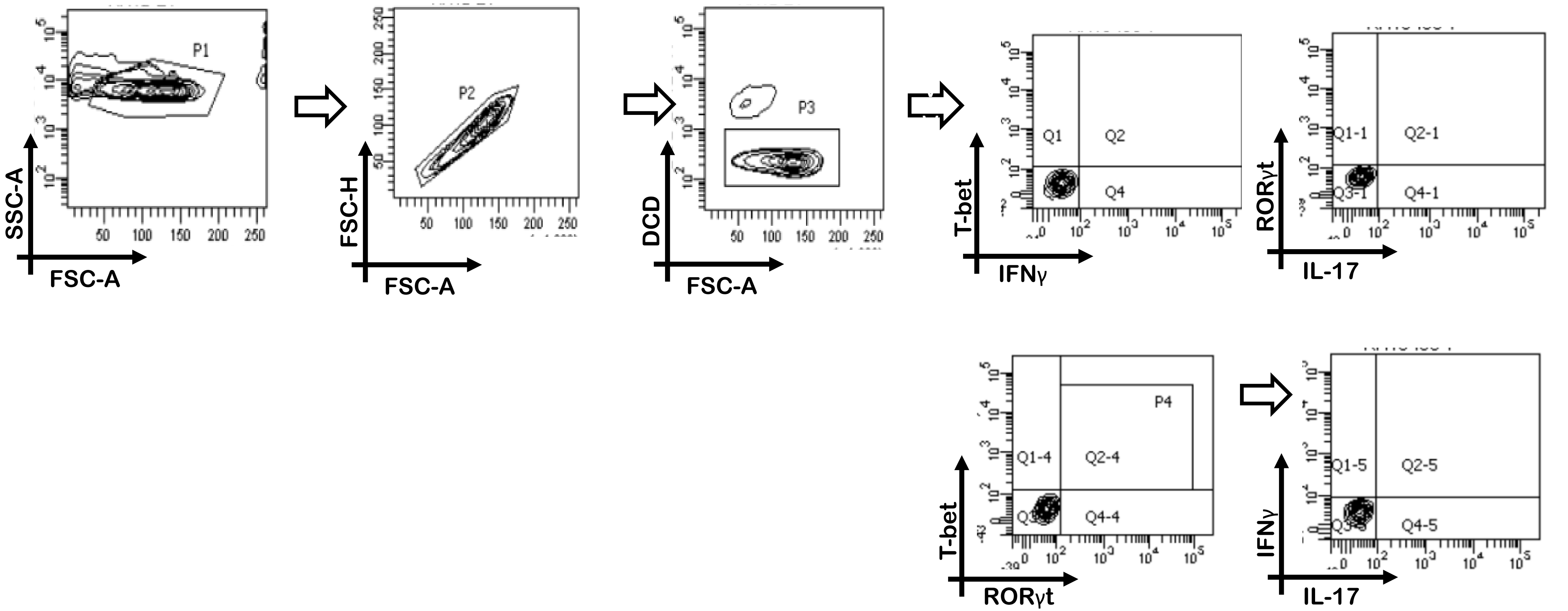


**Supplementary figure 2. Gating strategy for modulation in cytokine, transcription factor, T helper subsets expression upon hormone stimulation in RA CD4⁺ T cells.** Representative flow cytometry plots shows isolated CD4⁺ T cells where lymphocyte population is gated first, followed by singlet population. Live cells are gated using Zombie violet dye. The dual positive population of cytokines and transcription factor specific for Th1 cells (T-bet⁺ IFNγ⁺) and Th17 cells (RORγt⁺ IL-17⁺) are analyzed for any modulation.

**Supplementary Figure 3.**

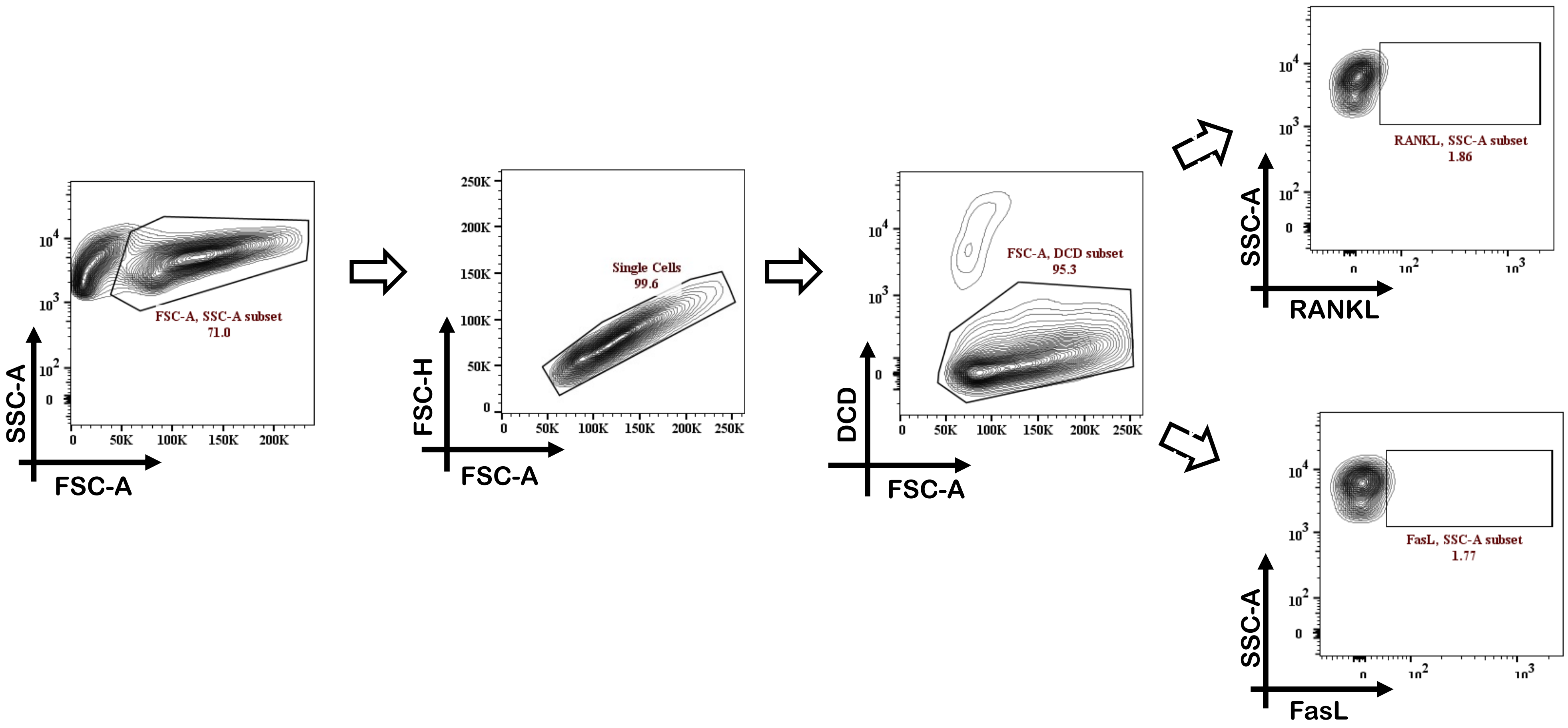


**Supplementary figure 3. Gating strategy for modulation in RANKL and FasL expression upon hormone stimulation in RA CD4⁺ T cells.** Representative flow cytometry plots shows isolated CD4⁺ T cells where lymphocyte population is gated first, followed by singlet population. Live cells are gated using Zombie violet dye. Modulation in RANKL and FasL was then analyzed.

**Supplementary Figure 4.**

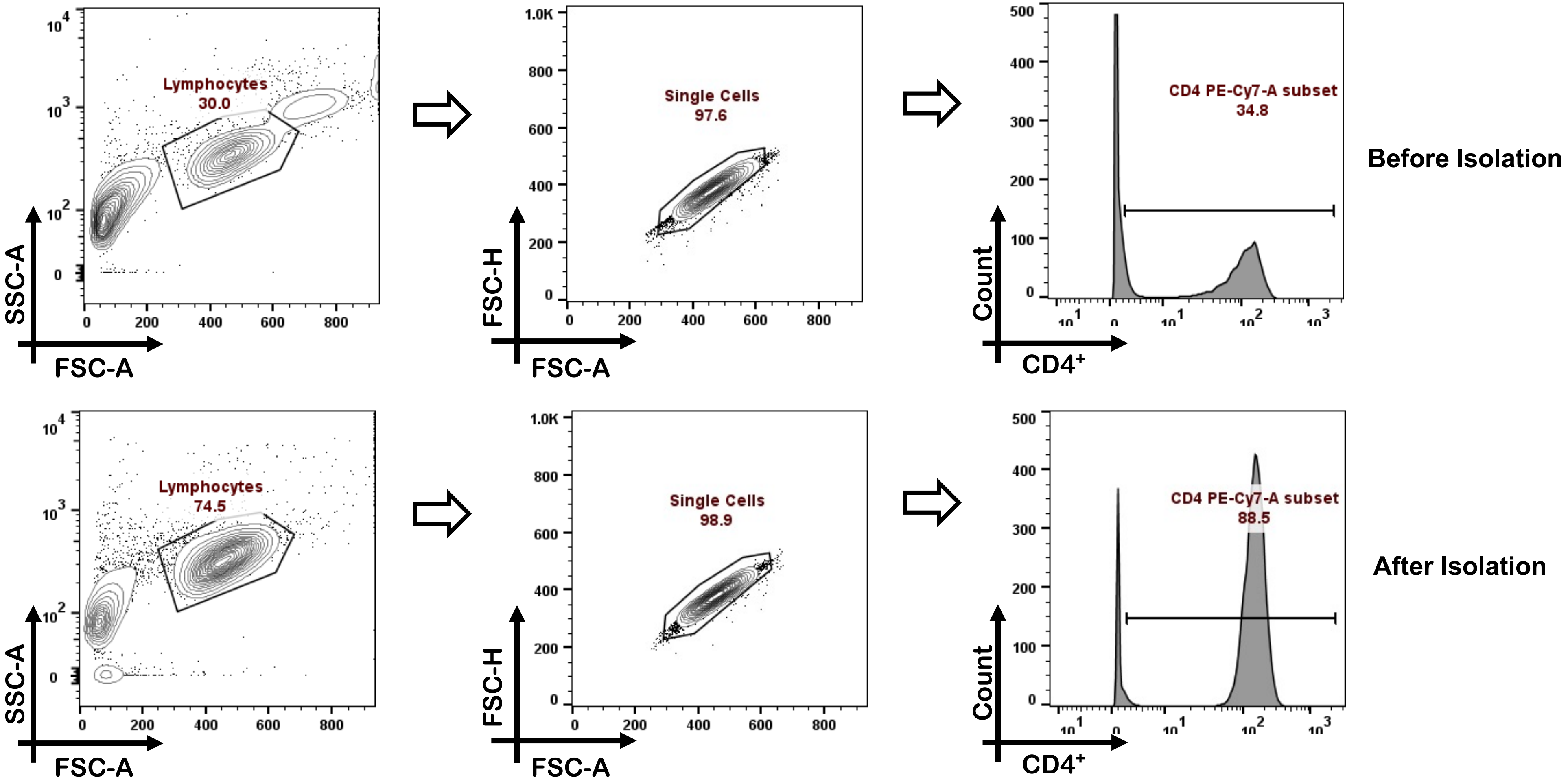


**Supplementary figure 4. CD4+ T cells purity validated prior *ex-vivo* experiments:** Representative flow cytometry plots shows purity of $CD4^{+}$ T cells isolated from healthy control PBMCs prior ex-vivo Th1 and Th17 differentiation studies

**Supplementary Figure 5.**

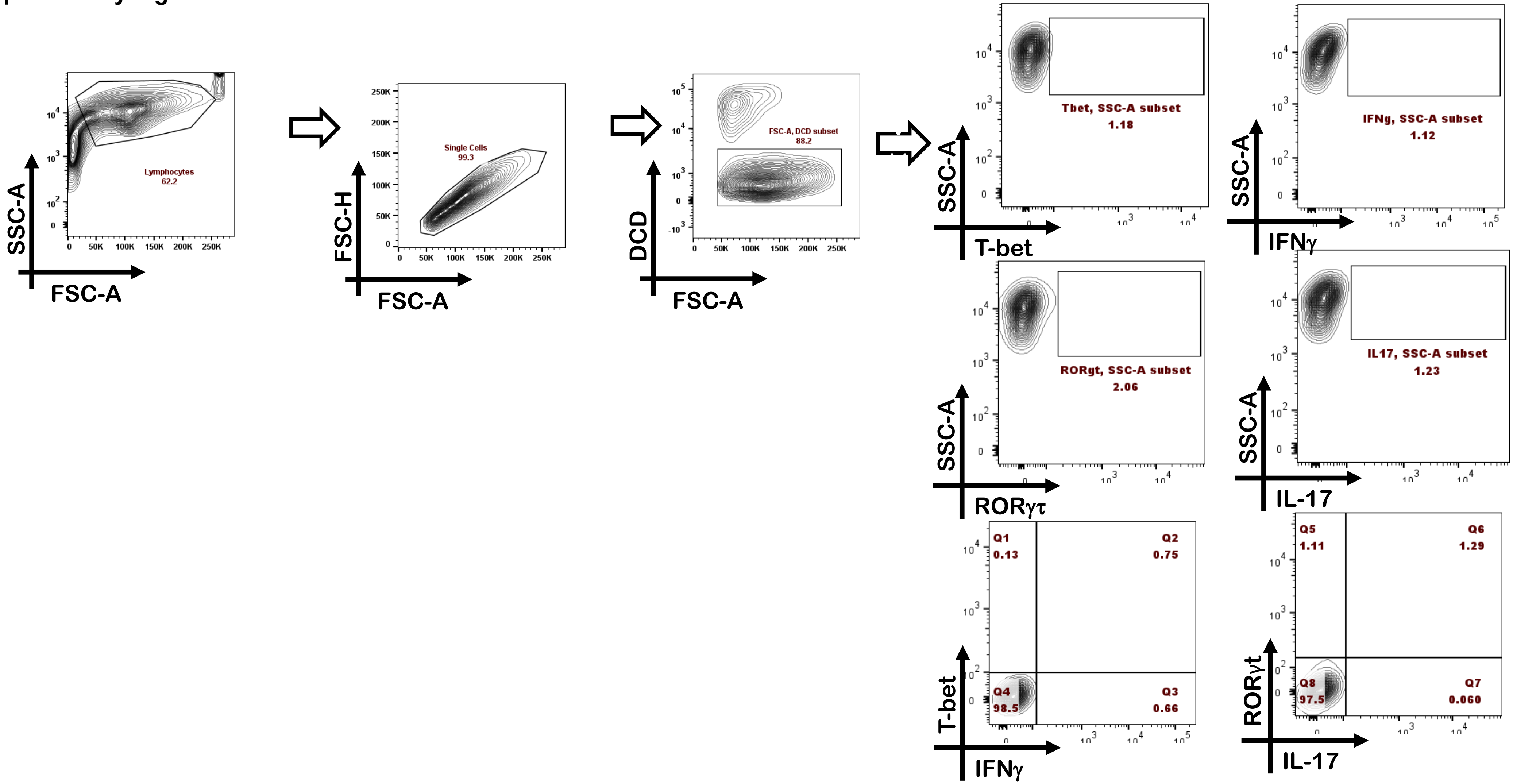


**Supplementary figure 5. Gating strategy for modulation in cytokine, transcription factor and T helper subset upon hormone stimulation in *ex-vivo* Th1 and Th17 cells.** Representative flow cytometry plots shows isolated CD4⁺ T cells where lymphocyte population is gated first, followed by singlet population. Live cells are gated using Zombie violet dye. Modulation in RANKL and FasL was then analyzed.

### Supplementary Figure 6.

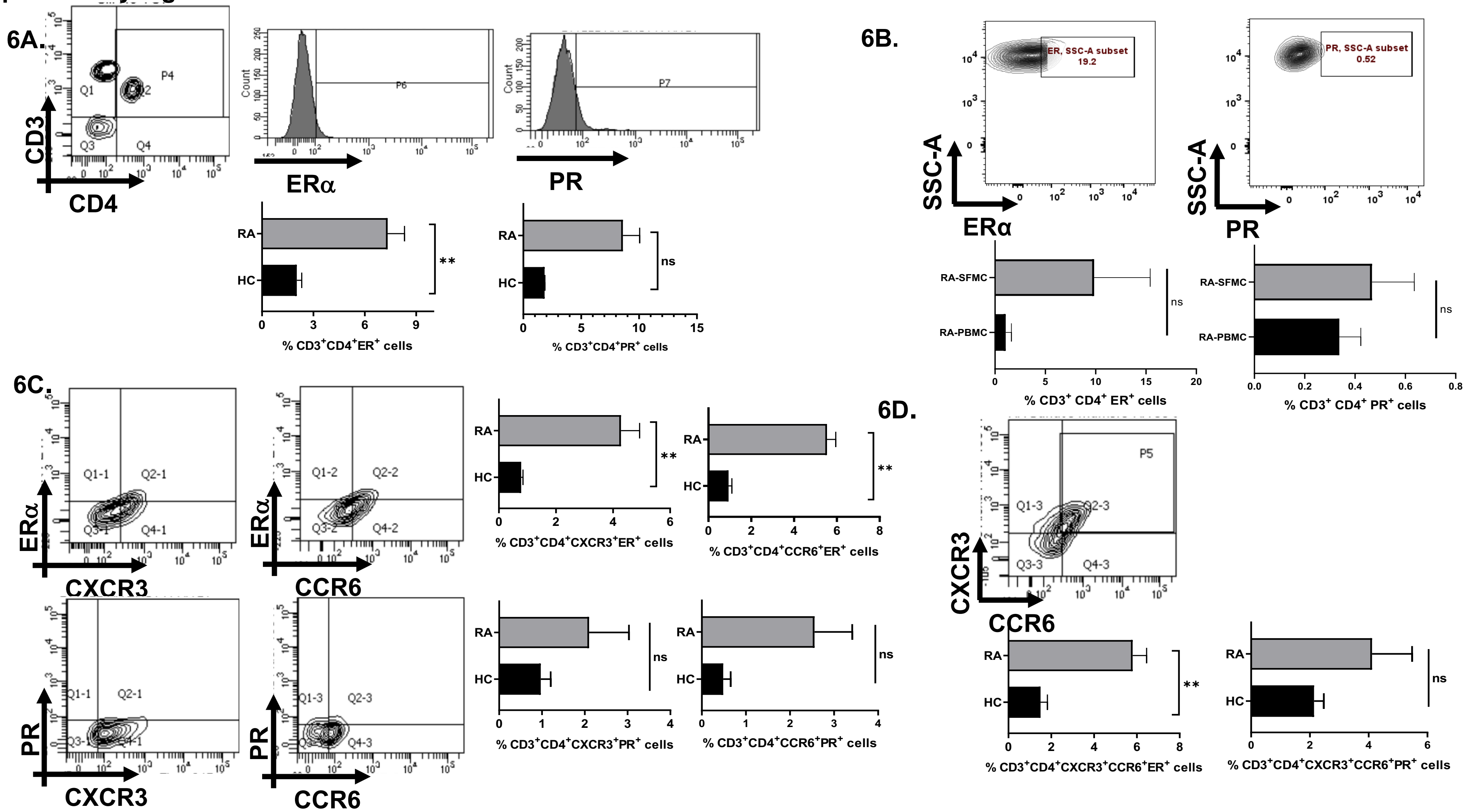


**Supplementary figure 6. Analysis of sex hormone receptors ERα and PR in CD4⁺ T cells of RA vs HC.** Representative flow cytometry, histogram and graphical plots shows differential expression of ERα and PR on CD4⁺ T cells,Th1 and Th17 subsets and dual positive population (Fig 6A,6C and 6D). Representative flow cytometry and graphical plots show insignificant differences in ERα and PR expression in paired RA samples (Fig-6B). Statistical analysis is based on Mann-Whitney U test to compare between the two groups.

**Supplementary Table 1. Reagents and software used in the study**

| S.no | Reagent | Catalogue no. |
|---|---|---|
| 1. | Human ProcartaPlex Mix & Match 46-Plex kit | Invitrogen<br>PPX-46-MX324DE |
| 2. | ProcartaPlex Human Antibody Isotyping Panels | Invitrogen<br>EPX070- 10818-901 |
| 3. | ProTM Human Cytokine Screening 48-Plex Panel | Bio-Rad<br>12007283 |
| 4. | Histopaque-1077 | Sigma<br>10771 |
| 5. | Hyaluronidase | Sigma Aldrich<br>H3506-100MG |
| 6. | Zombie fixable violet dye kit | BioLegend<br>423113 |
| 7. | RPMI 1640 | PAN-BIOTECH<br>P04-16520 |
| 8. | Fetal Bovine Serum | PAN-BIOTECH<br>P30-1402 |
| 9. | DPBS, w/o: Ca and Mg | PAN BIOTECH<br>P04-36500 |
| 10. | Dynabeads ™ Untouched Human CD4 T cell kit | Invitrogen<br>11346D |
| 11. | Phorbol 12-myristate 13-acetate (PMA) | Sigma<br>P8139-1MG |
| 12. | Ionomycin | Sigma<br>I0634-1MG |
| 13. | Brefeldin A (BFA) | Sigma<br>B6542-5MG |
| 14. | β-Estradiol powder | Sigma<br>E2758-1G |
| 15. | Progesterone powder | Sigma<br>P6149-1MG |
| 16. | BD Cytofix / Cytoperm ™ | BD Biosciences<br>554714 |
| 17. | eBiosciences™ FOXP3/ Transcription factor Staining Buffer Set | Invitrogen<br>00-5523-00 |
| 18. | FlowJO Version 10.8 | BD Biosciences |
| 19. | GraphPad Prism 9 | Dotmatics Pvt ltd. |

| S.no | Cytokines / Neutralizing antibodies | Catalogue No. |
|---|---|---|
| 1. | IL-12 Protein Human Recombinant | Thermo<br>200-12H-10UG |
| 2. | IL-2 Protein Human Recombinant | Prospec Bio<br>CYT-209 |
| 3. | IL-1β Protein Human Recombinant | Prospec Bio<br>CYT-208 |
| 4. | TGF-β Protein Human Recombinant | Prospec Bio<br>CYT-716 |
| 5. | IL-6 Protein Human Recombinant | Prospec Bio<br>CYT-213 |
| 6. | IL-21 Protein Human Recombinant | Prospec Bio<br>CYT-408 |
| 7. | IL-23 Protein Human Recombinant | Thermo<br>200-23-10UG |
| 8. | InVivoMAb anti-human CD3 | BioXcell<br>BE0001-2 |
| 9. | InVivoMAb anti-human/monkey CD28 | BioXcell<br>BE0291 |
| 10. | InVivoMAb anti-human IL-4 | BioXcell<br>BE0240 |
| 11. | InVivoMAb anti-human IFNγ | BioXcell<br>BEO235 |

| S.no. | Fluorochrome- tagged antibody | Catalogue No. | |
|---|---|---|---|
| | | **Antibody** | **Isotype** |
| 1. | CD3-AF 700 | Biolegend 317340 | Biolegend 400248 |
| 2. | CD4-Per CP Cy 5.5 | BioLegend 300530 | BioLegend 552834 |
| 3. | CD4- PECy7 | BioLegend 300512 | Invitrogen 25-4714-42 |
| 4. | CXCR3- AF488 | BioLegend 353710 | Invitrogen 53-4714-42 |
| 5. | CCR6- PECy7 | BD 560620 | Invitrogen 25-4714-42 |
| 6. | ERα-PE | CST 74244S | CST 5742S |
| 7. | PR A/B- AF 647 | CST 55652S | CST 2985S |
| 8. | IFNγ- AF 647 | BioLegend 502516 | BD 557783 |
| 9. | IL-17- PerCPCy5.5 | BD 560799 | BioLegend 552834 |
| 10. | TNF-α- AF700 | BD 557996 | BD 557882 |
| 11. | RORγt- PE | BD 563081 | BD 559529 |
| 12. | T-bet- PECy7 | Invitrogen 25-5825-82 | Invitrogen 25-4714-80 |
| 13. | RANKL- APC | BioLegend 347507 | BioLegend 982108 |
| 14. | FasL- BV711 | BD 744101 | BD 563044 |
